# RIPK1 Biallelic Activating Variants lead to Autoinflammatory Disease Driven by T cell Death

**DOI:** 10.1101/2024.03.28.24304774

**Authors:** Jialin Dai, Taijie Jin, Gaixiu Su, Xu Han, Jun Wang, Chenlu Liu, Wenjie Zheng, Qiuye Zhang, Jun Yang, Li Guo, Dan Zhang, Ming Li, Yingjie Xu, Tong Yue, Min Wen, Jia Zhu, Min Kang, Jianming Lai, Feifei Wu, Shihao Wang, Jiahui Zhang, Pui Y. Lee, Junying Yuan, Xiaomin Yu, Qing Zhou

## Abstract

Receptor-interacting serine/threonine-protein kinase 1 (RIPK1) is a key regulator of cell fate decision between pro-survival signaling and programmed cell death. The activation of RIPK1 is extensively modulated by posttranslational modification, such as ubiquitination. RIPK1 overactivation mediates autoinflammatory diseases in humans. However, the role of RIPK1 ubiquitination in human autoinflammatory diseases has not been reported, and the mechanism mediating the interaction of cell death and autoinflammation is largely unknown. Here, we report two patients carrying compound heterozygous *RIPK1* variants K377E/R390G with recurrent fevers, lymphadenopathy and skin rashes. Mechanistically, K377E and R390G mutations impaired ubiquitination of RIPK1, which suppresses the formation of TNFR1 signaling complex (TNF-RSC) and NF-κB activation, resulting in the loss of CD8^+^ T cells mediated by RIPK1 activation-induced cell death in the patients. We reveal that the death of CD8^+^ T cells promotes the secretion of TNF and IFNγ to activate monocytes and macrophages, which triggers further production of proinflammatory cytokines to amplify autoinflammation. Disruption of the communication between T cells and monocytes/macrophages through pharmacologic blockade of TNF and IFNγ attenuated proinflammatory cytokine production in macrophages. Collectively, our results demonstrate a crucial role of RIPK1 ubiquitination in regulating CD8^+^ T cell death and restraining autoinflammation. Our study demonstrates the mechanism for a group of autoinflammatory diseases mediated by RIPK1 activation-induced cell death, and highlights an important role of CD8^+^ T cells in driving autoinflammation.

## Introduction

Systemic autoinflammatory diseases (SAIDs) are a group of disorders induced by overactivation of innate immune system characterized by recurrent fever and multi-organ inflammation, but typically without high-titer antibodies or antigen-specific T cells. Distinct from autoimmune diseases defined by dysregulation of the adaptive immune system, SAIDs are believed to be mostly driven by the malfunction of innate immune cells such as monocytes and neutrophils ^1,2^. Therefore, the contribution of adaptive immune cells in triggering activation of myeloid cells has not been appreciated in the pathogenesis of SAIDs.

To date, more than 50 genes responsible for monogenic SAIDs have been identified. The list of genes whose mutations lead to SAIDs are enriched with key regulators of programmed cell death (PCD). Enhanced PCD is one of the characteristics in the SAID patients caused by the cleavage resistance mutations in *RIPK1*^3–5^ or deficiencies of *RIPK1*^6,7^, TANK-binding kinase 1 (*TBK1*)^8^, OTU deubiquitinase with linear linkage specificity (*OTULIN*), HOIL1-interacting protein (*HOIP*), Heme-oxidized IRP2 ubiquitin ligase-1 (*HOIL1*) and Shank-associated RH domain-interacting protein (*SHARPIN)*^9^. Pathogenic variants of these genes have been shown to activate apoptosis and necroptosis in cellular or murine models^10^. While excessive cell death is strongly linked to autoinflammatory diseases in humans, the mechanism by which cell death causes spontaneous inflammation in these conditions is unknown.

RIPK1 is a key mediator of cell death and inflammation^11^. Overactivation of RIPK1 leads to enhanced apoptosis, necroptosis and inflammatory response in mice and humans^12^. Whereas *RIPK1* deficiency and heterozygous non-cleavable variants in *RIPK1* cause immunodeficiency and dominantly inherited autoinflammatory diseases, respectively, a recessive autoinflammatory disease caused by *RIPK1* variants has not been reported. K63-linked ubiquitination of RIPK1 on K377 is essential for NF-κB pathway activation and negative regulation of cell death. Mice bearing *Ripk1*^K376R/K376R^ knockin mutation displays embryonic lethality due to excessive apoptosis and necroptosis caused by RIPK1 activation^13–15^. Ablation of *Fadd* and *Mlkl/Ripk3* rescues the embryonic lethality and allows them to grow into fertile adults. Nevertheless, the function of RIPK1 ubiquitination in human disease has not been defined and studied. In this study, we identified two autoinflammatory patients carrying recessively inherited compound heterozygous *RIPK1* variants K377E/R390G. We demonstrate that *RIPK1* K377E/R390G variants lead to autoinflammatory disease by prompting RIPK1 activation-mediated the death of CD8^+^ T cells.

## Results

### Biallelic *RIPK1* mutations in patients with autoinflammation

We identified two children from one family with autoinflammatory disease characterized by recurrent fevers, lymphadenopathy and skin rashes (Fig. 1a-d). The proband (Patient 1, P1) exhibited very early onset inflammatory bowel disease (VEO-IBD) characterized by diarrhea, abdominal pain and perianal abscess. At the age between 0-5, respectively, P1 and patient 2 (P2) displayed fever episodes that last 3-4 days and recurred every 8-9 days. Their fever episodes were accompanied by elevated serum C-reactive protein (CRP), serum amyloid A (SAA), erythrocyte sedimentation rate (ESR), D-dimer and lactate dehydrogenase (LDH) (Fig. 1e, Extended Data Fig. 1a-c). Neither case exhibited evidence of immunodeficiency or infection. In addition, the levels of pro-inflammatory cytokines IL-6 and TNF were strongly increased in the serum of both patients during flares (Fig. 1f). Serum CRP levels and adenosine deaminase (ADA) activity in patients were higher than that of unaffected controls and parents during disease flares and even in remission (Fig. 1f). P1 had taken prednisone during flares, with resolution of fever but limited effect on inflammatory indexes (Fig. 1e, Extended Data Fig. 1a-c).

**Fig. 1.**
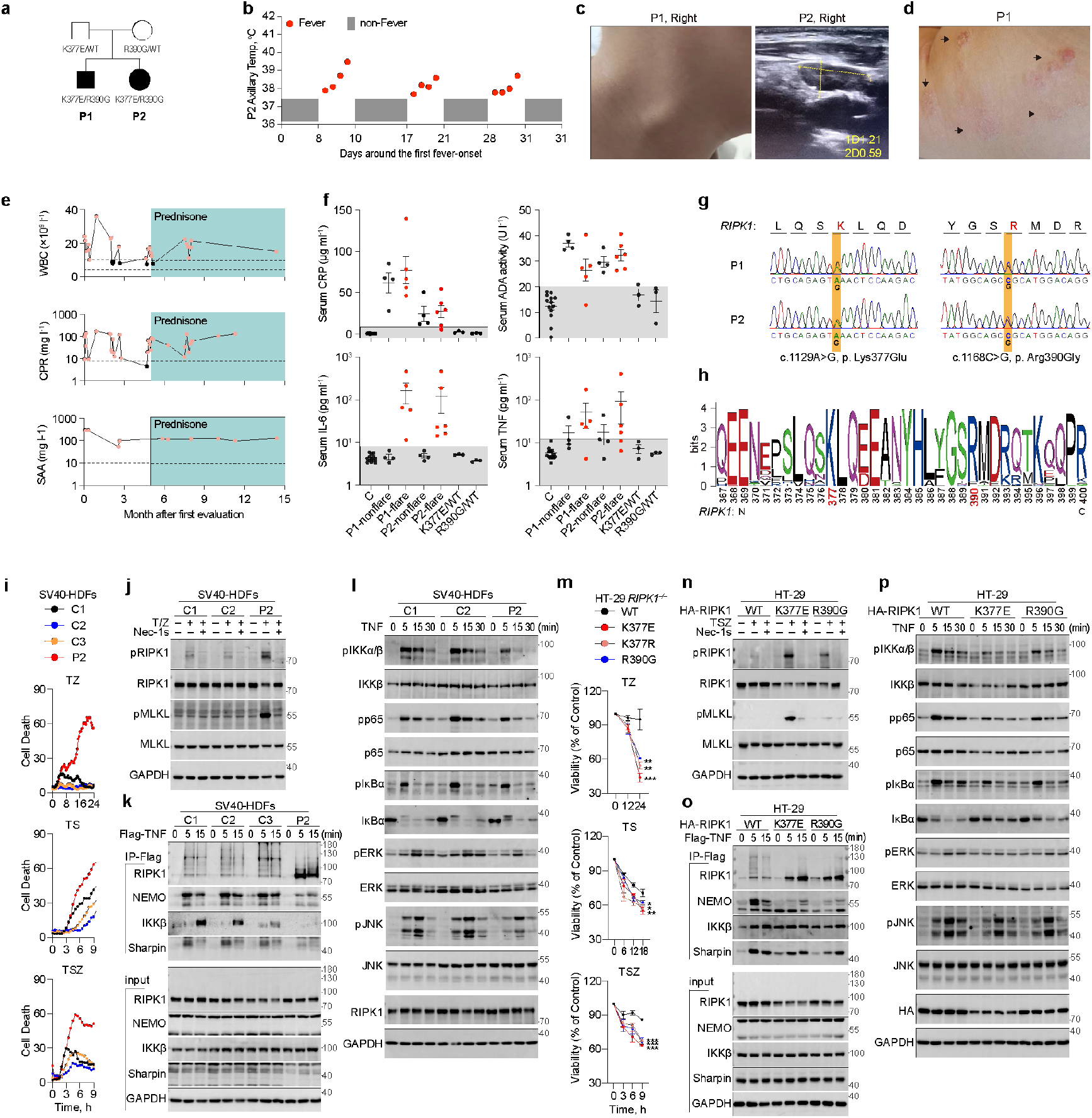
Compound heterozygous gain-of-function mutations in *RIPK1* causes systemic autoinflammatory diseases. **a**, Pedigrees of a family with two patients carrying mutations at *RIPK1* K377 and R390. Patient 1 (P1) and patient 2 (P2) were represented with filled symbols. **b,** Timeline of recurrent fever episodes in P2 over 5 weeks. Red dots represented increased temperatures during fever episodes and grey boxes represent the normal temperatures between flares. **c,** Representative sonographic image showing lymphadenopathy of P2. The diameters of right cervical lymph nodes were marked with dotted yellow line. **d,** Representative macroscopic image of dermatological manifestations in P1 during acute phase reaction, showing skin rashes (arrows). **e,** Whole blood cell counts (WBC), C-reactive protein (CRP) and serum amyloid A protein (SAA) were measured serially after the first evaluation of P1 and prednisone treatment (cyan shading) started at the age between 0-5. Horizontal dotted lines indicate age-specific high values for CRP, SAA or high and low values for WBC count. Red dots donate the abnormal values. **f,** C-reactive protein (CRP), serum ADA enzymatic activity and IL-6, TNF levels of unaffected controls (C, n=13), P1 during prednisone treatment (4 samples taken during flare and 5 samples taken during non-flare), P2 (4 samples taken during flare and 6 samples taken during non-flare), unaffected parents carrying heterozygous K377E mutation (K377E/WT, 1 sample from I-2 and 2 samples from II-2) or R390G mutation (R390G/WT, 3 samples from II-3). **g,** Sanger sequencing chromatograms of *RIPK1* showed heterozygous base substitutions at p. K377E and p. R390G in whole blood genomic DNA from P1 and P2. **h,** WebLogo demonstrating conservation of human *RIPK1* K377 and R390 across 42 vertebrate species. **i-l,** Cell death of SV40-immortalized human dermal fibroblasts (SV40-HDFs) isolated from three unaffected controls (C1-C3) and P2 were monitored by continuous imaging of SYTOX™ Orange staining. The SV40-HDFs were treated with T, S, Z as indicated (**i**). SV40-HDFs was treated with TZ with or without Nec-1s (10 μM) for 12 hours and immunoblotted with indicated antibodies (**j**). The TNFR signaling complex (TNF-RSC) were immunoprecipitated with Flag-TNF for 5 or 15 min and immunoblotted with indicated antibodies (**k**). NF-κB and MAPK activation of SV40-HDFs treated with TNF for 5, 15 or 30 min immunoblotted with indicated antibodies (**l**). **m-p,** *RIPK1*^-/-^ HT-29 cells were lentivirally complemented with HA-RIPK1 wild-type (WT), K377E, R390G and treated with T, S, Z as indicated. Cell viability was measured by CellTiter-Glo luminescent cell viability assay (**m**). The necroptosis markers (**n**), TNF-RSC (**o**) and the NF-κB and MAPK activation (**p**) were analyzed in the same way. T denotes 20 ng ml^-1^ TNF; S denotes 250 nM SM-164; Z denotes 25 μM z-VAD-fmk. Graphs show mean±SD. Statistical significance was determined by one-way ANOVA (**m**), **P < 0.01, ***P < 0.001.

By performing whole exome sequencing (WES) and sanger sequencing on the family, we identified that both P1 and P2 carried the *RIPK1* compound heterozygous mutations (c. 1129A>G / p. K377E and c. 1168C>G / p. R390G), whereas their unaffected mother and father were heterozygous carriers of *RIPK1* R390G and K377E, respectively (Fig. 1g, Extended Data Fig. 1d-e). Therefore, this autoinflammatory disease in the siblings appeared to be recessively inherited and segregated with the inheritance of both *RIPK1* variants. The lysine residue at amino acid position 377 is the site for K63-linked ubiquitination of RIPK1^16^, while the function of residue R390 has not been characterized. Both K377 and R390 residues are conserved in RIPK1 across species (Fig. 1h). The two *RIPK1* variants were predicted to be deleterious with their CADD scores well above that of the non-cleavable *RIPK1* mutations D324V, D324H and D324N (Extended Data Fig. 1f).

### RIPK1 activating mutations impair ubiquitination of RIPK1 and NF-κB activation and promote cell death

Given that RIPK1 is a widely expressed regulator of pro-survival NF-κB signaling and programmed cell death, we utilized SV40-immortalized human dermal fibroblasts (SV40-HDFs) derived from unaffected controls and P2 to assess the functional status of these signaling pathways. SV40-HDFs from P2 show increased sensitivity to apoptosis induced by TNF (T) with cIAP1/2 degrader SM-164 (S) as well as necroptosis induced by TNF (T) with pan-caspase inhibitor z-VAD-FMK (Z) or TSZ (Fig. 1i, Extended Data Fig. 2a), and enhanced RIPK1 activation as shown by phosphorylation at S166 and that of MLKL as shown by phosphorylation at S358, which were rescued by the RIPK1 kinase inhibitor Necrostatin-1s (Nec-1s) (Fig. 1j, Extended Data Fig. 2a). These results suggest that the compound R390G and K377E mutations in RIPK1 promote the activation of RIPK1.

**Fig. 2.**
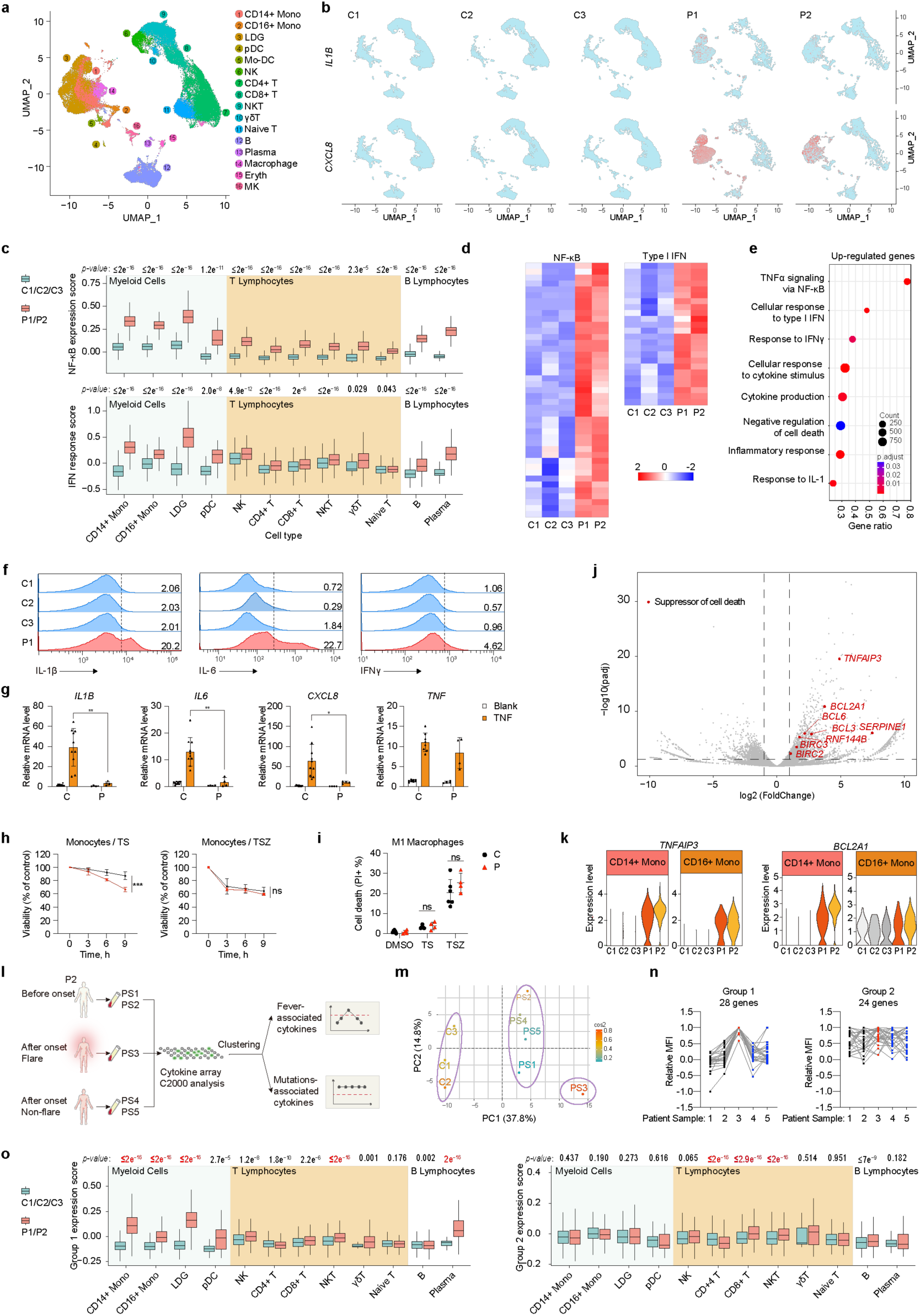
Strong activation of inflammatory signaling in myeloid cells carrying *RIPK1* mutations. **a**, Integrated uniform manifold approximation and projection (UMAP) visualization of peripheral blood mononuclear cells (PBMCs) from three unaffected controls (C1, n=13925 cells; C2, n=11117 cells; C3, n=12573 cells) and two patients (P1, n=10918 cells; P2, n=8516 cells), marker-based annotation of 15 cell subtypes were colored by cluster identity. LDG, low-density granulocyte; pDC, plasmacytoid dendritic cell; Mo-DC, monocyte-derived dendritic cell; NK, natural killer cell; NKT, natural killer T cell; Eryth, erythrocyte; MK, megakaryocyte. **b,** Visualization of expressions of *IL1B* and *CXCL8* (colored single cells) projecting PBMCs from unaffected controls (C1-C3) and two patients (P1 and P2) on UMAP plots. **c,** Boxplots of the NF-κB expression score (top panel) and IFN response score (bottom panel) of cell subtypes. **d,** Heatmap of representative genes downstream of NF-κB and type Ι IFN pathways in monocyte compartments. Gene names were listed in Extended Data Fig.4d. **e,** Dot plot showing the biological processes identified with un-regulated gene terms (FDR <0.05) in patients’ monocytes compared to that of unaffected controls using DAVID. The size of the circle indicates the gene count enriched in the pathway and the color represents pathway enrichment significance. **f,** Intracellular cytokine analysis of IL-1β, IL-6 and IFNγ in monocytes isolated from three unaffected (C1- C3) controls and P1. **g,** qRT-PCR analysis of *IL1B, IL6, CXCL8* and *TNF* mRNA levels in primary monocytes isolated from unaffected controls (n=8-10) and two patients (n=4 from P1 and P2) with or without 10 ng ml^-1^ TNF for 6 hours. **h,** CellTiter-Glo luminescent cell viability detection of primary monocytes isolated from unaffected controls (n=12) and two patients (n=3 from P1 and P2) stimulated with T, S, Z for indicated times. **i,** Statistics of dead M1 macrophages stimulated with TS or TSZ for 24 hours by imaging of propidium iodide (PI). M1 macrophages were differentiated from primary monocytes of unaffected controls (n=6) and two patients (n=4 from P1 and P2). Representative images were presented in Extended Data Fig.3e. **j,** Volcano plot displaying the differentially expressed genes in monocyte compartments between three unaffected controls (C1-C3) and two patients (P1 and P2). Representative suppressive genes of cell death were labeled with red dots and gene names. **k,** Clusters in the monocyte compartments showing the expression levels of *TNFAIP3* and *BCL2A1* in three unaffected controls (C1-C3) and two patients (P1 and P2). **l,** Schematic diagram showing serum cytokine microarray analysis of continuous P2’s serum samples before (PS1 and PS2) or after (PS3, flare; PS4 and PS5, non-flare) the onset of recurrent fever. **m,** PCA analysis of sample variance determined by semi-quantitative 174 cytokines antibody array on serum samples of C1-C3 from three unaffected controls and PS1-PS5 from P2. **n,** The integrated relative MFI plots of P2’s serum cytokine levels. 28 cytokines peaked in PS3 were enrich in group 1, 24 cytokines higher than control were enriched in group 2. **o,** Boxplots exhibiting mRNA expression scores of cytokines of group 1 (left panel) and group 2 (right panel) in indicated cell compartments. Group 1 and group 2 cytokine panels were clustered with the serum cytokine microarray analysis in Extended Data Fig.4. T denotes 10 ng ml^-1^ TNF; S denotes 250 nM SM164; Z denotes 25 μM z-VAD-fmk. All histogram graphs show mean±SD. Statistical significance was determined by two-tailed unpaired t-test (**c, g, i, l**) or one-way ANOVA (**h**), ns, *P* > 0.05, \**P*<0.05, \*\**P* < 0.01, \*\*\**P* < 0.001.

TNF-RSC is an important signaling complex that forms within minutes upon TNFR1 activation by TNF^17^. We next determined the effect of R390G and K377E compound mutations on TNF-RSC formation in SV40-HDFs upon stimulation with Flag-TNF. Ubiquitination of RIPK1 as well as recruitment of NEMO, IKKβ and SHARPIN to the TNF-RSC were impaired in P2 SV40-HDFs compared to unaffected controls (Fig. 1k). Furthermore, the activation of NF-κB and MΑPK signaling by TNF were impaired in P2 SV40-HDFs as revealed by reduced phosphorylation of IKKα/β, p65, IκBα, ERK and JNK (Fig. 1l).

The above findings were further investigated in *RIPK1*^-/-^ HT-29 cell lines complemented with wild-type (WT), K377E and R390G RIPK1. K377E and R390G compound mutations sensitized HT-29 cells to RIPK1-dependent cell death (Extended Data Fig. 2b-c). Furthermore, *RIPK1*^-/-^ HT-29 cells expressed K377E or R390G RIPK1 alone were also sensitized to TS and TSZ stimulation induced cell death to the same extent as that express both K377E and R390G (Fig. 1m-n, Extended Data Fig. 2d). Thus, K377E and R390G compound mutations or single mutation alone can promote cell death. Notably, the expression of K377E and R390G compound mutations or individual K377E and R390G mutation alone also reduced the levels of RIPK1 ubiquitination in TNF-RSC (Fig. 1o), as well as the activation of NF-κB and MΑPK (Fig. 1p), and the production of downstream cytokines (Extended Data Fig. 2e).

K377 is a ubiquitination site in RIPK1 intermediate domain (ID domain)^16^. Consistently, K377E, but not R390G, directly abolished the ubiquitination of ID domain (Extended Data Fig. 2f). Since R390 is not expected to be the site of ubiquitination, we considered the possibility that R390G might affect the recruitment of E3 ubiquitination complexes. Mind Bomb-2 (MIB2) is known to be an E3 ubiquitin ligase recruited to TNF-RSC to mediate RIPK1 ubiquitination^18^. We found that the interaction between Mind Bomb-2 (MIB2) and RIPK1 R390G was demonstrated to be significantly reduced, which partly explained for the impaired ubiquitination of R390G RIPK1 (Extended Data Fig. 2g). Collectively, these data demonstrate that both K377E and R390G mutations promote RIPK1 activation-dependent cell death and impaired pro-survival NF-κB signaling by modulating RIPK1 ubiquitination in TNF-RSC.

### Activation of inflammatory signaling in myeloid cells of the patients

We next studied the inflammatory signaling pathways by performing single-cell RNA sequencing on peripheral blood mononuclear cells (PBMCs) from P1, P2 and three unaffected controls. The monocyte compartment in patients was significant expanded (Fig. 2a, Extended Data Fig. 3a). Similar to the previously reported *RIPK1* D324V mutant^4^, transcriptional up-regulation of proinflammatory cytokines (*IL1B* and *CXCL8*) was detected in patient cells, especially in monocyte subsets (Fig. 2b). Likewise, differential gene expression analysis revealed greatest up-regulation of genes governed by NF-κB and type-I IFN pathways in myeloid cells from the patients (Fig. 2c-d, Extended Data Fig. 3b-c). The up-regulated genes in the patients’ monocytes were also enriched for pathways of cytokine production, cytokine stimulation, and negative regulation of cell death in the gene ontology (GO) analysis (Fig. 2e, Extended Data Fig. 3d), suggesting that myeloid cells play a crucial role in increased cytokine production and pro-inflammatory signal transduction in the patients. Increased basal expression of IL-1β, IL-6 as well as IFNγ in monocytes from P1 was confirmed by flow cytometry (Fig. 2f). However, monocytes isolated from the patients exhibited reduced transcription of NF-κB regulated genes including *IL1B*, *IL6*, *CXCL8* and *TNF* in response to TNF stimulation (Fig. 2g), in accordance with suppressed NF-κB activation in SV40-HDFs as demonstrated earlier (Fig. 1k, l). Thus, the increased inflammatory response of myeloid cells in the patients was likely represented a secondary effect triggered by K377E/R390G variants.

**Fig. 3.**
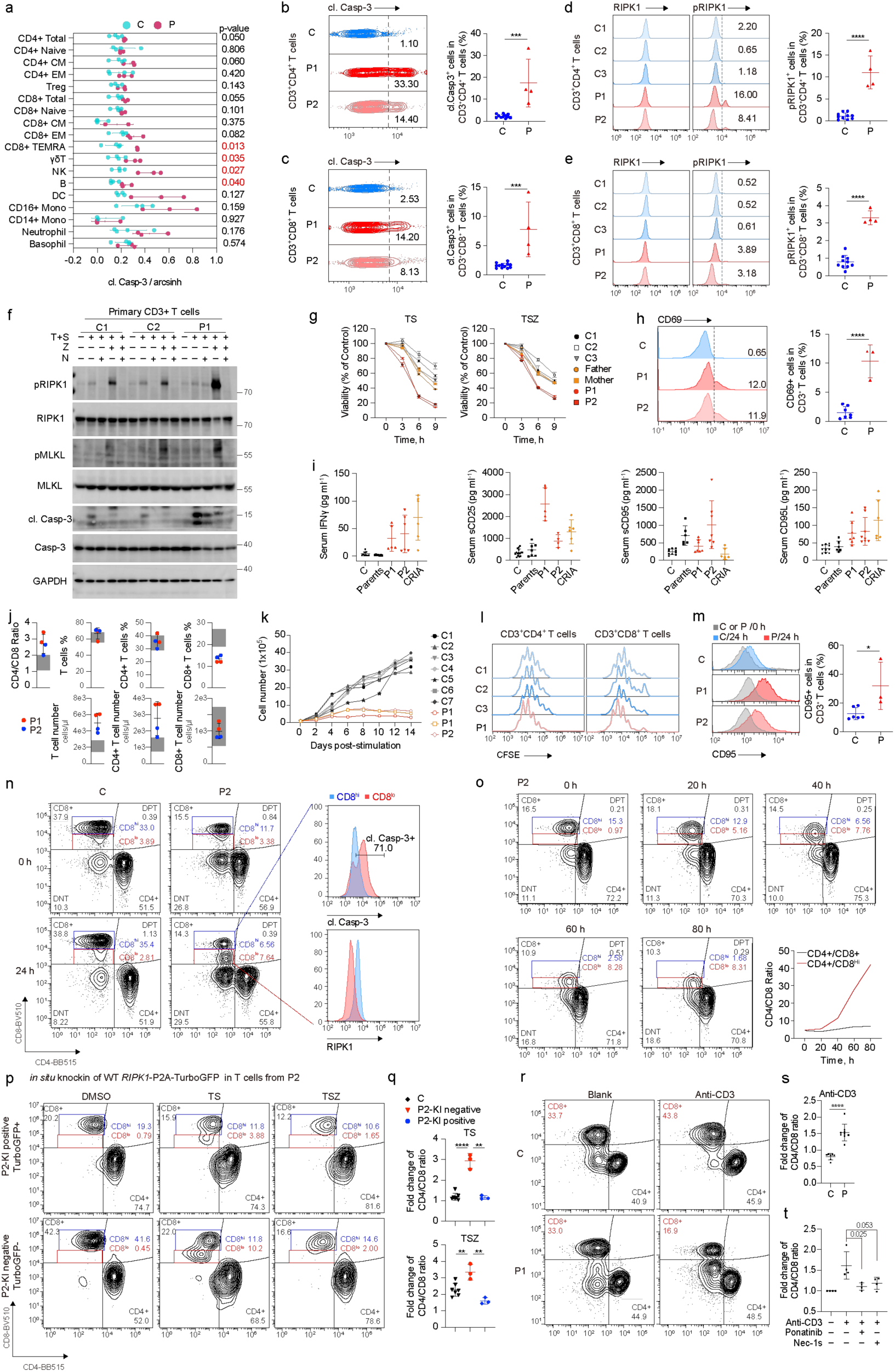
Increased cell death and CD4/CD8 ratio in T cells caused by RIPK1 activation. **a**, Average expression values (arcsinh transformed) of cl. Casp-3 across identified peripheral immune cell lineages in PBMCs freshly isolated from four unaffected controls (C) and two patients (P, 1 sample from P1 and 2 samples from P2) by mass cytometry by time-of-flight (CyTOF) analysis. **b-e,** Representative contour plots of cl. Casp-3 (**b-c**) and histograms of total RIPK1, pRIPK1(S166) (**d-e**) levels in CD4^+^ T and CD8^+^ T cells in freshly isolated PBMCs from unaffected controls (n=10-12) and patients (n=4 for P1 and P2) (left panel). Summary histograms of cl. Casp-3 and pRIPK1(S166) (right panel) were shown. **f,** Immunoblotting analysis of primary CD3^+^ T cells isolated from two unaffected controls (C1 and C2) and P1 treated with T and S with or without Z, N for 6 hours. Antibodies as indicated were immunoblotted to show the activation of apoptosis or necroptosis. **g,** CellTiter-Glo luminescent cell viability detection of primary CD3^+^ T cells isolated from three unaffected controls (C1-C3), two patients (P1 and P2) and their parents treated with TS or TSZ for indicated times, n=4. **h,** Representative histogram and statistical analysis of surface CD69 expression on CD3^+^ T cells from unaffected controls (n=7) and two patients (n=3 for P1 and P2). **i,** Serum IFNγ, sCD25, sCD95 and CD95L of unaffected controls (C, n=10), two patients (P1, n=5-7; P2, n=4-8) and their parents (n=6-7) as well as two CRIA syndrome patients (n=6) determined by ELISA. **j,** CD4/CD8 ratio in T cells, percentages and absolute counts of CD3^+^ T, CD4^+^ T, CD8^+^ T cells in whole blood sample of P1 (red dots) and P2 (blue dots) measured using immunophenotyping. **k,** *In vitro* expansion of primary T cells isolated from unaffected controls (n=7) and patients (n=3 for P1 and P2) as indicated. Cell numbers were counted every two days post anti-CD3/CD28 dynabeads stimulation. **l,** Proliferation of CD4^+^ T and CD8^+^ T cells isolated from three unaffected controls (C1-C3) and P1 were analyzed using CFSE staining upon stimulation with anti-CD3/CD28 dynabeads for 96 hours. **m,** Representative histograms of surface CD95 expression on CD3^+^ T cells analyzed with flow cytometry after culturing *in vitro* for 24 hours. **n,** Representative contour plots depicting the percentages of CD4^+^ T, CD8^+^ T, double negative T (DNT) and double positive T (DPT) in CD3^+^ T cells gated from PBMCs of P2 culturing *in vitro* for 24 hours. Histograms displaying cl. Casp-3 and RIPK1 expression levels in CD8^hi^ T and CD8^lo^ T cells. **o,** Representative contour plots and summary histograms showing percentages of CD4^+^ T, CD8^+^ T and CD8^hi^ T cells and CD4/CD8 ratio in CD3^+^ T cells isolated from P2 cultured for indicated times. **p-q,** Representative contour plots showing percentages of CD4^+^ T, CD8^+^ T in CD3^+^ T cells of P2 with (TurboGFP^+^) or without (TurboGFP^-^) restoration using CRISPR/Cas9-mediated *in situ* knock-in of WT *RIPK1-P2A-TurboGFP* under stimulation of T with or without S, Z for 6 hours (**p**). CD4/CD8 ratio in CD3^+^ T cells of unaffected controls (n=7) and two patients (n=3 for P1 and P2) were quantified (**q**). **r-s,** Representative contour plots (**r**) and statistical analysis (**s**) showing percentages of CD4^+^ T, CD8^+^ T, DNT and DPT cell in CD3^+^ T cells isolated from unaffected control (n=7) and two patients (n=9 for P1 and P2) under the stimulation of anti-CD3 antibody (5 μg ml^-1^) for 72 hours. **t,** Statistical analysis of fold change of CD4 /CD8 ratio in CD3^+^ T cells isolated from two patients (n=4 for P1 and P2) under stimulation of anti-CD3 antibody (5 μg ml^-1^) with or without 250 nM Ponatinib or 10 μM Nec-1s for 24 hours. For representative contour plots, see Extended Data Fig. 6h. T denotes 10 ng ml^-1^ TNF; S denotes 250 nM SM164; Z denotes 25 μM z-VAD-fmk; N denotes 10 μM Nec-1s. All histogram graphs show mean±SD. Statistical significance was determined by one-way ANOVA (**g)** or two-tailed unpaired t-test in the rest, \**P*<0.05, \*\**P* < 0.01, \*\*\**P* < 0.001, \*\*\*\**P* < 0.0001.

As RIPK1 overactivation can promote programmed cell death leading to inflammation^19^, we assessed the cell death response of monocytes to TNF stimulation. The monocytes from the patients were more sensitive to TS-induced apoptosis than that of unaffected controls, whereas they showed similar sensitivity to necroptosis induced by TSZ (Fig. 2h). In addition, no differences of apoptosis and necroptosis were observed between monocyte-derived macrophages (MDMs) from patients and that from unaffected controls (Fig. 2i, Extended Data Fig. 3e). On the other hand, although the expression of cell death suppressor genes *TNFAIP3*, *BCL2A1*, *BCL6*, *BCL3*, *RNF144B*, *BIRC2* and *BIRC3* were elevated in patients’ various immune cell types, monocytes, LDG and DC cells express these genes significantly higher than that in lymphocyte subsets (Fig. 2j, k and Extended Data Fig. 3f). These results suggest that the elevated inflammatory response of monocytes induced by K377E/R390G variants may be regulated non-autonomously by other cell types that were more susceptible to cell death.

To define the upstream factors that drove the overproduction of inflammatory cytokines in monocytes, we employed semi-quantitative protein microarray analysis on serum samples from unaffected controls and P2 (Fig. 2l-m). We collected samples before and after the first fever episode (disease onset) for P2. Differentially expressed cytokines were clustered into three groups according to the data trend. Twenty-eight cytokines in group 1 (such as IL-1β) peaked during flare, reflecting a direct correlation with fever status (Fig. 2n, Extended Data Fig. 4a-b). 24 cytokines in group 2 (such as sCD95) (Fig. 2n, Extended Data Fig. 4c-d) and 32 cytokines in group 3 (Extended Data Fig. 4e-f) continuously changed across samples of P2. Up-regulated cytokines in group 2 were considered to be directly regulated by the *RIPK1* K377E/R390G mutations.

**Fig. 4.**
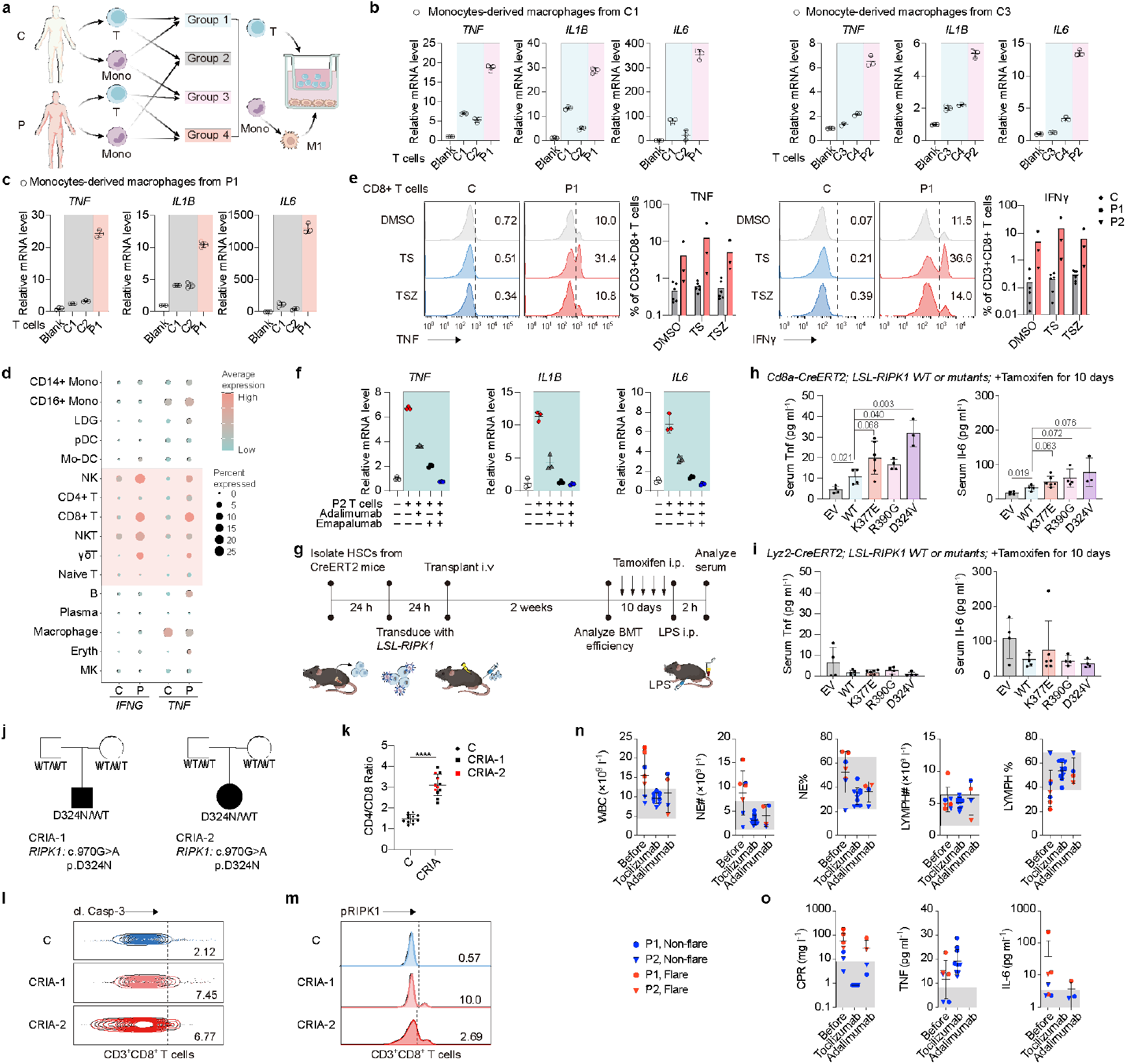
RIPK1 activation causes inflammation through T-Mono axis. **a**, Schematic diagram of T cell-MDM co-culture system. **b-c,** qRT-PCR analysis of *TNF, IL1B* and *IL6* mRNA levels in GM-CSF-activated monocytes derived macrophages (MDMs) co-cultured with or without T cells for 6 hours. GM-CSF-activated MDMs generated from unaffected control 1 (C1) (left panel) or unaffected control 3 (C3) (right panel) (**b**) or P1 (**c**) were co-cultured with donor-matched T cells as well as T cells from other unaffected controls (C2 and C4) or patients as indicated. **d,** Dot plot of *IFNG, TNF* in cell subtypes found in scRNA-seq of PBMCs from three unaffected controls and two patients. The size of the circle indicates the percentage of cells positive for gene expression and the color represents the average expression level. **e,** Representative histograms and statistical analysis of TNF and IFNγ expression in CD8^+^ T cells in PBMCs isolated from unaffected controls (n=6) and patients (n=3 for P1 and P2) treated with 10 ng ml^-1^ TNF (T) with or without 250 nM SM-164 (S), 25 μM z-VAD-fmk (Z) for 24 hours. **f,** T cells isolated from P2 were co-cultured with MDMs generated from unaffected control in the presence of 10 μg ml^-1^ adalimumab, 10 μg ml^-1^ emapalumab alone or in combination for 6 hours. *TNF, IL1B* and *IL6* mRNA levels of MDMs were analyzed by qRT-PCR. **g,** Experimental timeline for generation of HSCs-transplanted mice with conditional overexpression of human WT or mutated RIPK1 K377E, R390G or D324V in CD8α^+^ T cell- or myeloid cell-specific lineage. **h-i,** ELISA measurement of serum Tnf and Il6 levels in mice specifically overexpressing WT or mutated hRIPK1 in CD8α^+^ T cells (**h**) or myeloid cells (**i**). **j-m,** Pedigrees of two families with CRIA patients (CRIA-1 and CRIA-2) (**j**). Patients were represented with filled symbols. Summary histograms of CD4/CD8 ratio in CD3^+^ T cells of unaffected controls (n=12), CRIA-1 (n=7) and CRIA-2 (n=5) were shown (**k).** Representative contour plots of cl. Casp-3 (**l**) and histograms of pRIPK1(S166) (**m**) protein levels in CD8^+^ T cells in freshly isolated PBMCs from CRIA patients. **n-o,** Whole blood cell counts (WBC), cell counts and proportions of neutrophil (NE#-NE%) and lymphocyte (LYMPH# -LYMPH%) (**n)**, Serum C-reactive protein (CRP), TNF and IL-6 (**o**) of P1 (red dots) and P2 (blue dots) measured before and after tocilizumab or adalimumab treatments. The grey area represents the range of unaffected controls. Dots represent each sample and graphs show mean±SD. Statistical significance was determined by two-tailed unpaired t-test, \**P*<0.05, \*\**P* < 0.01, \*\*\**P* < 0.001, \*\*\*\**P* < 0.0001.

To further define the source of the cytokines in group 1 and 2, we compared the expression scores of the two groups between patients and unaffected controls by analyzing single cell RNA-seq data. The cytokines in group 1 were most significantly up-regulated in patients’ monocytes, low-density granulocyte (LDG), plasmacytoid dendritic cell (pDC) and plasma cells (Fig. 2o), consistent with the strong inflammatory signature in myeloid cells during fever. Notably, the genes encoding cytokines in group 2 were up-regulated most significantly in the patients’ lymphocyte subsets, especially in CD8^+^ T cells (Fig. 2o), implying a pathogenic role of K377E/R390G variants in T lymphocytes. In addition, the expression levels of cell death suppressor genes in T lymphocytes were also lower compared with monocytes, LDG, pDC and plasma cells (Extended Data Fig. 3f), suggesting that T lymphocytes may be sensitized to undergo programmed cell death. Collectively, these results supported a critical role of monocytes in promoting inflammation and implied that T lymphocyte subsets may also contribute to the pathophysiology of this autoinflammatory disease.

### RIPK1 activation leads to increased cell death and CD4/CD8 ratio in T cells

To better understand the impact of *RIPK1* mutations on immune profile, we performed cytometry by time-of-flight (CyTOF) on PBMCs isolated from unaffected controls and patients. CyTOF result revealed that CD4^+^T, B and γδ T cell proportions in patients were elevated whereas CD8^+^ T and NK cell proportions were reduced comparing to unaffected controls (Extended Data Fig. 5a-b). Apoptosis indicated by cleaved caspase-3 was increased in CD8^+^ TEMRA, γδT, NK and B cell subsets of patients (Fig. 3a). Flow cytometric analysis confirmed that patients’ CD4^+^ and CD8^+^ T cells, not CD19^+^ B cells and CD14^+^ monocytes, displayed augmented activation of RIPK1 and cleaved caspase-3 (Fig. 3b-e, Extended Data Fig. 5c-f). Furthermore, CD3^+^ T cells isolated from patients were more sensitive to TS-induced apoptosis and TSZ-induced necroptosis. They displayed higher levels of RIPK1 phosphorylation at S166 and MLKL phosphorylation at S358 comparing to that of unaffected controls, and these patterns were inhibited by treatment with Nec-1s (Fig. 3f-g), indicating that T cells bearing K377E/R390G mutations were more prone to RIPK1-dependent cell death. In accordance with the recessive inheritance, no significant differences of cell death were observed between unaffected parents carrying heterozygous mutation and unaffected controls.

The CyTOF results also revealed a global activation of patients’ immune cells as indicated by CD69 expression, while elevated CD38 expression appeared to be restricted to B cells and T cells (Extended Data Fig. 5g-h). Increased expression of CD69 in T cells was further validated by flow cytometry (Fig. 3h). Concomitantly, higher levels of T cell activation-related proinflammatory mediators including sCD25, TNF and IFNγ were observed in the serum of P1 and P2 as well as (cleavage-resistant RIPK1-induced autoinflammatory) CRIA patients (Fig. 3i). Activated lymphocytes undergo cell expansion and indeed we observed an elevation in the number of total lymphocytes, as well as that of CD3^+^ T cells, CD4^+^ T cells and B cells, except for that of cytotoxic CD8^+^ T cells and NKT cells in the immunophenotyping results (Fig. 3j, Extended Data Fig. 6a). As a consequence of the loss of CD8^+^ T cells, CD4/CD8 ratio was significantly increased in patients. *In vitro* expansion of T cells isolated from patients upon anti-CD3/CD28 stimulation were abnormal, but the proliferation ability was undampened (Fig. 3k-l), reflecting the occurrence of T cell death. Specifically, CD8^+^ T cells went through a contraction phase after activation presumably resulted from activation-induced cell death (AICD) ^20^. The augmented signal of CD95 (Fas) on the surface of patients’ T cells accompanied by elevations of serum sCD95 and CD95L levels also supported increased AICD (Fig. 3i, m).

To further characterize the abnormalities in the patient T cells, we analyzed T cell subsets from *in vitro* cultured PBMCs. We observed that CD8 expression was decreased in approximate half of the CD8^+^ T cells after culturing PBMCs isolated from P2 *in vitro* for 24 hours, while the expression on CD8^+^ T cells of unaffected controls remained normal (Fig. 3n). Through gating on CD8^hi^ (CD8^+^ T cells with normal CD8 expression) or CD8^lo^ (CD8^+^ T cells with lower CD8 expression) T cells, we found that in contrast to CD8^hi^ T cells, most of the CD8^lo^ T cells exhibited augmented cleaved caspase-3 (Fig. 3n), indicative of activated apoptosis. In addition, total RIPK1 level in CD8^lo^ T cells decreased which may be caused by caspase-3 cleavage (Fig. 3n). The decreased CD8 expression in patients’ T cells and the increases in CD4/CD8 ratio were more prominent after prolonged *in vitro* culture and after TS or TSZ stimulation compared to that of control (Fig. 3o, Extended Data Fig. 6b-d). The elevation of CD4/CD8 ratio induced by TS or TSZ could be inhibited by RIPK1 kinase inhibitor or *in situ* knockin of WT *RIPK1* (Fig. 3p-q, Extended Data Fig. 6e-f). In contrast, neither ferroptosis induced by RSL3 nor RIPK1-independent apoptosis induced by staurosporine (STS) can differentiate cell death of CD8^+^ T cells from the patients compared to that of unaffected controls (Extended Data Fig. 6g). In addition, the activation of patients’ T cells by anti-CD3 antibody stimulation increased CD4/CD8 ratio, which could be reversed by ponatinib or partially reversed by Nec-1s (Fig. 3r-t). Since ponatinib inhibits the kinase activity of RIPK1 and RIPK3 ^21^, these results suggest that they might play important roles in regulating AICD in the patient T cells bearing K377E/R390G compound RIPK1 mutations. Together, these results suggested that T cells, especially CD8^+^ T cells, were the most susceptible hematopoietic lineage to RIPK1-mediated cell death in patients with the *RIPK1* K377E/R390G variants.

### RIPK1 activation-induced T cell death drives inflammation in myeloid cells

Given the differential impact of the *RIPK1* K377E/R390G variants on T cells and monocytes, we next investigated cell-cell communication between these two cell types using a transwell system (Fig. 4a). The results showed that patients’ T cells induced higher cytokine expression in MDMs comparing to that of T cells isolated from unaffected controls (Fig. 4b, c). However, MDMs from patients showed less response than that of unaffected controls upon culturing with the identical T cells. The impaired inflammatory response by patients’ MDMs may be explained by the suppressed NF-κB activation caused by *RIPK1* K377E/R390G mutations. Interestingly, patients’ T cells could still trigger massive cytokine production in autologous MDMs (Extended Data Fig. 7a). Thus, we conclude that increased cell death by T cells from the patients can act to potentiate the inflammatory response of monocytes and macrophages with *RIPK1* K377E/R390G mutations.

Previous studies have proposed TNF and IFNγ secreted by chimeric antigen receptor (CAR) T cells as key contributors to the activation of macrophage during cytokine release syndrome (CRS) ^22^. TNF combined with IFNγ were also reported to be a key inducer of inflammation in patients with severe COVID-19 ^23^. Intriguingly, IFNγ and TNF were specifically upregulated in patients’ T lymphocytes and NK cells whereas receptors of these two cytokines were highly expressed in myeloid cells (Fig. 4d, Extended Data Fig. 7b, c), providing additional support for T cell-monocyte/macrophage communication through these two receptor-ligand pairs. In comparison to unaffected controls, T cells, especially CD8^+^ T cells from patients, exhibited greater production of IFNγ and TNF. The production of these cytokines was further enhanced upon TS or TSZ stimulation (Fig. 4e, Extended Data Fig. 7d). These results indicated that IFNγ and TNF produced by activated T cells may be the key factors to initiate the inflammatory response of MDMs. Consistently, increased production of *TNF*, *IL1B* and *IL6* in MDMs generated from unaffected controls stimulated by either patient T cells or anti-CD3/CD28-activated donor-matched T cells could be partially inhibited by neutralization of IFNγ or TNF alone. The cytokine production was completely blocked by combined blockade of IFNγ and TNF (Fig. 4f, Extended Data Fig. 7e). Collectively, these data demonstrated that RIPK1 overactivation in CD8^+^ T cells promoted cell death as well as TNF and IFNγ secretion to trigger inflammatory cytokine production in MDMs (Extended Data Fig. 7f).

In order to validate this model and assess whether the function of *RIPK1* K377E and R390G mutations in CD8^+^ T cells was sufficient to induce inflammation, we exploited the Cre-loxp system combined with bone marrow transplantation to conditionally express WT or mutated human RIPK1 in specific hematopoietic compartments (Fig. 4g, Extended Data Fig. 8a). HSCs isolated from donor *Cd8a*-creERT2 mice or *Lyz2*-creERT2 mice were transduced with lentivirus encoding *EF1α*-promoted *loxp-GFP-stop-loxp-TagBFP2-P2A-human RIPK1* expression cassettes (*LSL-hRIPK1*). Afterwards, transduced HSCs were transplanted into lethally irradiated donor mice followed by tamoxifen induction. Transplantation efficiency and the specificities of *Cd8a* and *Lyz2* promoters were validated (Extended Data Fig. 8b-h). GFP expression in HSCs reflected transduction efficiency, and TagBFP2 confirmed WT/mutated RIPK1 expression in specific cell types upon tamoxifen induction (Extended Data Fig. 9a-d). We then evaluated the serum levels of inflammatory cytokines in the recipients. Under basal conditions, serum Tnf and Il-6 levels were higher in mice with Cd8α^+^ T cells expressing mutated RIPK1 (K377E, R390G or D324V) than WT RIPK1 and empty vector (EV) controls (Fig. 4h). No significant differences were observed between mice bearing myeloid cells overexpressing EV/WT RIPK1 controls and mutated RIPK1 (Fig. 4i). The result demonstrated the pathogenic function of K377E and R390G as well as the initiator role of T cells in the autoinflammation. We also investigated the function of *RIPK1* K377E/R390G mutations in myeloid cells. As TLR4 is specially highly expressed in myeloid cells, we assessed the responsiveness of recipients to LPS at a nonlethal dose. The results showed that LPS challenge led to elevation of Il-6 and Tnf in all groups. Recipients transplanted with HSCs expressing mutated RIPK1 (K377E, R390G or D324V) under the control of *Lyz2*, but not *Cd8a* promoter displayed higher serum level of inflammatory cytokines than WT and EV controls under LPS stimulation (Extended Data Fig. 9e-f), which is consistent with myeloid-specific inactivation of TAK1 that caused Ripk1 activation and cell death in mouse^24^.

*RIPK1* mutations at D324 ^3,4^, which could activate RIPK1-dependent cell death, have been reported to cause CRIA syndrome, respectively. Therefore, we examined the CD4/CD8 ratio and RIPK1 activation in CRIA patients (CRIA-1 and CRIA-2) to verify our model. Similar to patients with *RIPK1* K377E/R390G variants, we observed elevated CD4/CD8 ratio paired with increased RIPK1 phosphorylation and cleaved caspase-3 in PBMCs from CRIA patients (Fig. 4j-m, Extended Data Fig. 10a-b), indicating that CD8^+^ T cell death might be a general phenomenon during the inflammation of CRIA. Notably, *in vitro* culture also induced decreased CD8 expression in CD8^+^ T cells of CRIA-1 (Extended Data Fig. 10c). CD4 and CD8 expression levels in CRIA patients’ T cells were validated by CyTOF and CD8 expression decreased more in CD8^+^ TEMRA cells than other subsets, reflecting higher levels of cell death in CD8^+^ TEMRA cells (Extended Data Fig. 10d-g).

### Targeted therapies

As TNF and IFNγ were considered to mediate the inflammatory cytokine production of myeloid cells, we compared the treatment effectiveness of TNF inhibitor adalimumab with tocilizumab which blocks the signaling of IL-6 mainly produced by myeloid cells in patients (Fig. 4n-o). Both P1 and P2 responded to tocilizumab and showed marked alleviation of the elevated temperature and skin rashes and the inflammatory indexes including abnormal CRP, WBC, neutrophil and lymphocyte. However, the serum levels of TNF in both patients could not be suppressed by tocilizumab. TNF-targeting therapy is projected to partially interrupt the communication between T cells and monocytes/macrophages in our model. P2 responded to anti-TNF therapy with reduced severity of the symptoms and decreased frequency of fevers. However, adalimumab had limited effects on WBC and lymphocyte counts as well as symptoms of P1 as adalimumab alone was not enough to suppress serum levels of IL-6 (Fig. 4o), in accordance with the partial inhibitory effect of adalimumab on IL-6 produced by MDMs in the co-culture assay.

## Discussion

This is the first report of biallelic mutations K377E/R390G in *RIPK1* leading to early-onset SAIDs with manifestations of periodic fever, intermittent lymphadenopathy, skin rashes and VEO-IBD. Mutation at K377 blocks K63-linked ubiquitination of RIPK1, while function of *RIPK1* R390 has not been reported. Here, we revealed that both K377E and R390G mutations lead to defects in TNF-RSC formation and activation of RIPK1, which promote cell death and inflammation. Since K377E and R390G are the recessive gain-of-function (GOF) mutations of *RIPK1*, as are the previously described caspase cleavage resistance variants, we termed these two conditions as “Gain-of-function *RIPK1* variants-induced autoinflammatory syndrome (GRIA)”.

The condition in our study is inherited in an autosomal recessive (AR) pattern, which is different from autosomal dominantly inherited CRIA syndrome even if they share similar manifestations. Mutation in *RIPK1* D324 does not impair NF-κB pathway^3–5^; in contrast, compound heterozygous mutations in *RIPK1* K377/R390 suppressed NF-κB activation, indicating that the autoinflammation caused by these mutations is not directly induced by dysregulation of NF-κB. The evidence that PBMCs and MEFs carrying heterozygous mutations in D324 as well as T cells carrying compound heterozygous variants in *RIPK1* K377/R390 instead of heterozygous variant are hypersensitive to TNF-induced cell death support the inheritance pattern of the two conditions^3,4^. Thus, excessive RIPK1-mediated cell death triggered by heterozygous D324 mutations or biallelic K377E and R390G mutations is supposed to be the inducer of autoinflammation. Mutations of genes such as *TBK1*^8^, *OTULIN*, *HOIP*, *HOIL1* and *SHARPIN*^9^, haven been shown to cause several SAIDs. Variants of these genes have been studied in murine models and are revealed to cause embryonic lethality attributed by enhanced cell death^17,25–27^. In contrast, patients with programmed cell death-driven SAIDs (PCD-driven SAIDs) only manifest periodic inflammation with excessive production of proinflammatory cytokines such as IL-6 and TNF. Overactivation of cell death has been observed in the cells derived from these patients, supporting an important role of cell death in the pathogenesis of SAIDs. However, how programmed cell death induces autoinflammation in human diseases remains uninvestigated.

Our study provides a mechanism that K377E/R390G variants of *RIPK1* induce CD8^+^ T cell death to cause SAIDs. CD8^+^ T cells bearing K377E/R390G have the propensity to be activated and expanded, and then enter the contraction phase induced by programmed cell death. During this process, CD8^+^ T cells secret massive TNF and IFNγ which prime monocytes or macrophages to produce proinflammatory cytokines. The accumulation of the inflammatory cytokines such as group 2 cytokines secreted from monocytes would induce the onset of fever. At the same time, inflammation accelerates the proliferation of T lymphocytes and autoinflammation with a positive feedback loop that amplifies the inflammatory reaction (Extended Data Fig. 7f). Eventually, inflammation is terminated with the decrease of activated CD8^+^ T cells. Thus, we hypothesize that dynamics of CD4/CD8 ratio is correlated with the inflammatory state. Consistently, we captured once the resting stage before T cell expansion of P2 as well as CRIA-2 with the normal ranges of WBC, lymphocyte counts and CD4/CD8 ratio. Therefore, increased CD4/CD8 ratio combined with augmented RIPK1 activation in T cells are supposed to be biomarkers for PCD-driven inflammation. This evidence for diagnosis is verified in patients with CRIA syndrome. But it is worth noting that intensive gating strategy is required as CD8^lo^ T cells representing the dying CD8^+^ T cells may be considered as CD8^+^ T cells by conventional gating strategy, leading to the falsely normal CD4/CD8 ratio. Since RIPK1 activation-induced AICD of T cells trigger the inflammation in SAID patients carrying *RIPK1* GOF variants, RIPK1 kinase inhibitors or RIPK1 proteolysis targeting chimeras degraders may provide a promising strategy for the treatment of SAID induced by RIPK1 overactivation^11^.

GRIA syndrome is characterized by selective cell death of CD8^+^ T cells as reflected by the elevation of CD4/CD8 ratio in peripheral lymphocytes. This process is remarkably similar to the contraction phase of CD8^+^ T cells following infections as most effector CD8^+^ T cells undergo programmed cell death after clonal expansion upon antigen-induced activation. Interestingly, intermittent fever with viremia has been shown to be correlated with CD4/CD8 ratio, which is similar to high CD4/CD8 ratio in K377E/R390G patients during flare^28^. Although the two diseases have completely different mechanism, it is worthwhile to note that almost 14 days were needed for CD8^+^ T cells to undergo expansion and contraction upon antigen stimulation^29^, which is identical to the period of intermittent fever in GRIA patients.

Taken together, our findings provide a new insight that not only innate immune system, but also adaptive immune cells can play a causative role in the onset of SAIDs. We demonstrate that myeloid cells, such as monocytes and macrophages, act as executors of PCD-driven SAIDs for that they take responsibility to produce excessive cytokines to induce autoinflammation. This finding supports the notion that SAIDs is induced by dysregulation of innate immune system. In addition, we also highlight the essential role of T cells in PCD-driven SAIDs as they are the most sensitive hematopoietic compartments to cell death and has the predisposition to undergo AICD which motivate inflammatory response of myeloid cells, indicating the importance of cell-cell communication in pathogenesis of PCD-driven SAIDs. Hence, blockade of communication between T cells and myeloid cells could be a promising therapeutic strategy for PCD-driven SAIDs.

## Acknowledgements

We thank the patients, their families and the unaffected controls for their support during this research study. We also thank the core facility of the Life Sciences Institute, Zhejiang University, and core facilities of Liangzhu Laboratory, Zhejiang University. The works of Q.Z. were supported by grants 82225022, 32141004 and 32321002 from the National Natural Science Foundation of China. The works of X.Y. were supported by the Hundred-Talent Program of Zhejiang University. J.D. received grant 32300618 from the National Natural Science Foundation of China. J.W. received the grant 2023M733104 from China Postdoctoral Science Foundation. L.G. received the grant LHDMY23H100005 from Zhejiang Provincial Natural Science Foundation of China.

## Author contributions

Q.Z., X.Y., and J.Y. directed and supervised the research. J.D., T.J., and G.S. contributed equally. J.D. and T.J. designed the study, performed experiments and analyzed most of the data. X.H., J.W., and C.L. analyzed data. S.W. and J.Z. performed experiments. G.S., W.Z., Q.Z., J.Y., P.Y.L., D.Z., M.L., Y.X., T.Y., M.W., J.Z., M.K., and J.L. enrolled the patients, collected and interpreted clinical information. J.D., T.J., Q.Z., J.Y., and P.Y.L. wrote the manuscript, with input from others. All authors contributed to the review and approval of the manuscript.

## Methods

### Patients

Patients (P1 and P2; CRIA-1 and CRIA-2) were evaluated under protocols approved by Institutional Review Board and Ethical Committee at the Children’s Hospital of Zhejiang University School of Medicine (protocol 2021-IRB-172). P1 and P2 were evaluated at Children’s Hospital affiliated to the Capital Institute of Pediatrics. All relevant ethical regulations were followed. All patients provided written informed consent.

### Whole exome sequencing and genotyping with sanger sequencing

Whole blood genomic DNA was extracted using Maxwell RSC Whole Blood DNA Kit (Promega, AS1520) and 1 μg DNA was used for whole-exome sequencing. WES and data analysis were performed as previously described^4^. For genotyping of other affected or unaffected family members, 437 bp genomic DNA fragment containing *RIPK1* K377/R390 was amplified with forward primer 5’-TTGAGA TACAGATTCATGACGGCG-3’ and reverse primer 5’-CTGGTGCACTGCATTACCATGAC-3’ for sanger sequencing. For genotyping of CRIA patients, 514 bp genomic DNA fragment containing *RIPK1* D324 was amplified with forward primer 5’-AAATCAGGAAGTGTGAGTCCTACAACC-3’ and reverse primer 5’-TGTTCAGAACACTCTATGACTGGTGAG-3’ for sanger sequencing.

### Single-cell RNA sequencing

PBMCs of patient 1 and patient 2 were freshly isolated and 10000 single-cell were captured by 10X Genomics Chromium for cDNA library preparation according to the manufacturer’s instruction. Sequencing was carried out on Illumina NovaSeq PE150 platform by Novogene Company (Beijing, China). The resulting count matrices followed the standard pipeline with default parameters. Further data analysis was performed as previously described ^4^.

### Cell lines, cell isolation, culture and stimulation

The HEK293T cell line was from the American Type Culture Collection (ATCC). Human PBMC samples were generated from peripheral blood samples using gradient centrifuging with LSM™ lymphocyte separation medium (MP Bio, Cat.50494X). Then primary T lymphocytes and monocytes were isolated from PBMCs by negative selection using Pan T Cell Isolation Kit (Miltenyi Biotec, 130-096-535) and Pan Monocyte Isolation Kit (Miltenyi Biotec, 130-096-537) according to manufacturer’s instructions. SV40-HDFs were derived from skin biopsies of patient or control donors as described and transformed by lentiviral transduction of SV40-large T antigen.

The HEK293T cells, SV40-HDFs were cultured in Dulbecco’s Modified Eagle Medium (DMEM, Gibco) supplemented with 10% FBS. Human PBMCs, primary T cells and monocytes were cultured in RPMI 1640 (Gibco) supplemented with 10% FBS. HT-29 cells were cultured in McCoy’s 5A medium (Gibco) supplemented with 10% FBS.

Recombinant human TNF (Peprotech, 300-01A) was used to stimulate HT-29 cells (20 ng ml^-1^), SV40- HDFs (20 ng ml^-1^) and human primary monocytes (50 ng ml^-1^) for the indicated times. z-VAD-fmk (Selleck, S7023, 25 μM), SM-164 (Selleck, S7089, 250 nM) and Nec-1s (Selleck, S8641, 10 μM) were used to treat HT-29 cells, human primary T cells and monocytes. Staurosporine (MCE, HY-15141, 100 nM) and RSL3 (Selleck, S8155, 1 μM) were used to induce apoptosis or ferroptosis in T cells.

### Flow cytometry

Cells were washed with FACS buffer (0.5% BSA in PBS) and blocked with human Fc Block (BD Biosciences, 564219) followed by staining with antibodies against surface molecules in dark for 30 min at 4 °C. Then cells were washed twice and stained with Fixable Viability Stain 780 (BD Biosciences, 565388) to distinguish live versus dead cell. For intracellular staining, cells were fixed and permeabilized with BD Phosflow™ Fix Buffer and Perm Buffer (BD Biosciences: 557870, 558052) and then washed twice and stained with antibodies against intracellular antigens for 30 min at 4 °C. For pRIPK1 (Ser166) and cleaved caspase-3 staining, cells were incubated with Alexa Fluor 647 labeled goat anti-rabbit IgG (Thermo, A-21244) for 30 min at 4°C after primary antibody staining. Cells were acquired on CytoFlex (Beckman) and data were analyzed using FlowJo software. The following antibody clones were used for staining in flow cytometry analysis: CD3 (SK7, BD Biosciences), CD4 (RPA-T4, BD Biosciences), CD8 (RPA-T8, BD Biosciences), CD14 (M5E2, BD Biosciences), CD16 (3G8, BD Biosciences), CD19 (SJ25C1, BD Biosciences), CD56 (NCAM16.2, BD Biosciences), IL-1β (JK1B-1, BioLegend), TNF (MAb11, BD Biosciences), IL-6 (MQ2-13A5, BD Biosciences), IFNγ (4S.B3, BD Biosciences), Cleaved Caspase-3 (Asp175) (CST, #9661), pRIPK1(Ser166) (D8I3A, CST), RIPK1 (D94C12, CST).

### RNA isolation and real-time RT-PCR

Total RNA from cells was extracted using a RNeasy Mini Kit (Qiagen, 74104) according to the manufacturer’s instructions. 1 μg RNA was reverse transcribed into cDNA with ReverTra Ace® qPCR RT Master Mix (TOYOBO, FSQ-301). cDNA was then diluted and used for real-time PCR using 2x Universal SYBR Green Fast qPCR Mix (Abclonal, RK21203) with primers targeting *IL1B* (F, 5’- TGCTCTGGGATTCTCTTCAGC-3’; R, 5’-AAGTCATCCTCATTGCCACTGT-3’), *IL6* (F, 5’- CATCCTCGACGGCATCTCAG-3’; R, 5’-ACCAGGCAAGTCTCCTCATTG-3’), *IL8* (F 5’- CCAAACCTTTCCACCCCAAA-3’; R, 5’-GAATTCTCAGCCCTCTTCAAAAAC-3’) *TNF* (F, 5’- CTCTTCTGCCTGCTGCACTTTG-3’; R, 5’-ATGGGCTACAGGCTTGTCACTC-3’), and *GAPDH* (F, 5’-TCGACAGTCAGCCGCATCTTC-3’; R, 5’-GCGCCCAATACGACCAAATCC-3’). Relative transcription levels were calculated by normalization to *GAPDH* mRNA.

### Cell viability assay and cell death assay

Primary T cells (15000 cells/well), SV40-HDFs (2000 cells/well) or HT-29 (5000 cells/well) were plated in 384-well plate and were stimulated with indicated conditions. General cell viability was measured by using CellTiter-Glo® Luminescent Cell Viability Assay (Promega, G7572) according to the manufacturer’s instructions. The percentage of viability was normalized to untreated controls. Cell death detection was performed by direct staining with 0.5 μg/ml Propidium Iodide (PI) and 0.5 μg/ml DAPI for 10 min and imaging with fluorescence microscope. PI positive dead cell was counted and normalized to DAPI counts. For continuous cell death measurement, cells were simulated with indicated conditions and stained with 200 nM Hoechst 33342 (Beyotime, C1022) and 200 nM SYTOX™ Orange (Thermo, S34861) at the same time. The percentage of dead cell was assayed every 1 hour by imaging with BioTek Cytation 3 for 18 hours with 5% CO2 and 37 °C condition. Results were exported as counts per well to be processed and graphed using Prism 9.

### Protein array

Semiquantitative analysis of 174 cytokine levels of P2’s serum samples before (normal stage) or after (onset, non-flare) the first onset of fever was performed with human Cytokine Array G2000 (RayBiotech) by Novogene Company (Beijing, China). Generally, the cytokine array glass slides were firstly blocked for 1 hour at room temperature and then incubated with 100 μl serum samples at 4 °C overnight. On the second day, the slides were washed with wash buffer and incubated with Biotin-Conjugated Anti-Cytokines and HiLyte Plus™ 555 Streptavidin-Fluor according to the manufacturer’s instructions. The results were scanned with InnoScan 300 Microarray Scanner (Innopsys) at 532 nm with 10 μm resolution. The mean fluorescence intensity (MFI) was calculated and exported for further analysis.

### Human and mouse cytokines measurement

Human or mouse serum cytokines were measured by enzyme-linked immunosorbent assay (ELISA) (Multi Sciences: 70-EK1280, 70-EK194, 70-EK182HS, 70-EK106HS, 70-EK191, 70-EK180HS, 70-EK1F01, 70-EK1F02, EK206HS, EK282HS/2) according to the manufacturer’s instructions. T cell culture supernatant cytokines were determined by Cytometric Bead Arrays (BD Biosciences) according to the manufacturer’s instructions. Human serum adenosine deaminase (ADA) activity was measured with Boyle colorimetric method by using ADA Activity Assay Kit (Yuanye Bio, R22207) according to the manufacturer’s instructions. Student’s t-test was performed for the statistical analysis.

### *In vitro* cell proliferation assay

Fresh isolated CD3^+^ T cells were labeled with CFSE (Thermo, 65-0850-84, 5 μM) and then stimulated with anti-CD3/CD28 beads (T&L Biotechnology) at 1:1 ratio supplemented with hrIL-7 (PrimeGene) and hrIL-15 (PrimeGene) in 48-well plates for 96 hours. Then all cells were co-stained with CD4-BB515 and CD8-BV510 antibodies and analyzed using CytoFlex (Beckman).

### Gene editing of patient T cells

Primary T cells isolated from patients were stimulated with anti-CD3/CD28 beads for 3 days. For electroporation, 180 pmol sgRNA and 25 μg Cas9 protein were mixed and incubated for 15 min at room temperature for ribonucleoprotein complexes (RNP) formation. Then 3e6 T cells resuspended in opti-MEM were mixed with RNP and 6 μg dsDNA template and electroporated using Gemini X2 (BTX) under the #905 program for human primary T cells. After electroporation, 1 ml warm X-VIVOTM 15 Serum-free Hematopoietic Cell Medium (Lonza) supplemented with 10% FBS was immediately added to the cuvette and the mixture was then carefully transferred to 24-well plates for further culture and analysis.

### T cell-MDM co-culture assay

Freshly isolated monocytes were seeded in 24-well plate and differentiated into monocyte-derived macrophages (MDMs) with 20 ng/ml GM-CSF treatment for 10 days. Then, 0.4 µm pore polycarbonate membrane inserts containing T cells isolated from PBMCs of patient or healthy donors were placed into the 24-well plate for the T cell-MDM co-culture at the ratio of 5:1 for 8 hours. Finally, T cells were removed and bottom MDMs were collected for mRNA extraction.

### Mice

All mouse studies complied with relevant ethical regulations and approved by Institutional Animal Care and Use Committee (IACUC) of the Zhejiang University (Registration No.:27043). The *Cd8a*-creERT2 and *Lyz2*-creERT2 strains on C57BL/6J background were purchased from GemPharmatech. The *Rosa26*- mTmG strain on C57BL/6J background was purchased from Cyagen. All mice were maintained under specific pathogen-free condition with free access to food and water at Laboratory Animal Center of Zhejiang University.

### HSC isolation, transduction, transplantation, induction and LPS challenge

Mouse hematopoietic stem cells (HSC) were isolated from 6- to 8-week-old donor mice using Lineage cell depletion kit (Miltenyi Biotec, 130-090-858) according to manufacturer’s instruction. HSCs were plated in 12-well plate and were transduced with lentivirus carrying human RIPK1 inducible expression system (MOI=50). Transduction efficiencies were measured with flow cytometry after 24 hours culturing and 200,000 of transduced HSCs were transplanted into lethally irradiated (4 Gy, 2 times) recipient mice (female, 8 to 12 weeks old). Transplantation efficiencies were validated by flow cytometry analysis after 2 weeks. Successfully transplanted mice were injected i.p with 20 mg/ml tamoxifen (MCE, HY-13757A) dissolved in corn oil (MCE, HY-Y1888) every two days for 5 times to induce the expression of WT, K377E, R390G or D324V mutated RIPK1. Then, these mice were injected i.p with 0.5 mg/kg lipopolysaccharide (LPS) (Sigma, L6529) and serum were collected before injection or injected for 2 hours. Serum Il6 and Tnf were measured by ELISA as described.

### Immunoprecipitation and western blot

For TNFR signaling complex (TNF-RSC) enrichment, 5e6 cells were seeded and treated with flag tagged TNF (ENZO, ALX-522-008) for indicated times. The samples were immediately washed with cold PBS and lysed in cold lysis buffer (25 mM Tris-HCl, pH 7.4, 150 mM NaCl, 0.5% NP-40, 10% glycerol) with freshly added inhibitors blocking protease and phosphatase (Merck: 4906837001, 4693159001) for 30 min. Then samples were cleared through centrifugation at 20000 RCF for 10 min and protein concentrations were determined by Pierce™ BCA Protein Assay Kits (Thermo, 23225). Then the TNF-RSC was immunoprecipitated with FLAG M2 magnetic beads (Sigma, M8823) at 4 °C for 4 hours and the protein complex was directly eluted by boiling in 1×SDS loading buffer. For TNF downstream signaling analysis, 4e5 cells were seeded and treated as indicated and then cells were harvested and lysed directly with sample loading buffer. These samples were separated through routine SDS-PAGE and transferred to 0.45 μm nitrocellulose membrane (Cytiva, 10600002) and blotted with indicated antibodies.

### CyTOF

PBMCs were freshly isolated from whole bloods of four patients (P1 and P2; CRIA-1 and CRIA-2) and four unaffected controls, then blocked and labeled with conjugated antibodies as described^31^. Fixed cell samples were resuspend analyzed on a mass cytometer (Helios, Fluidigm). The CyTOF data exported as FCS files were gated in FlowJo (v10.0.7) to remove doublets and CD45^+ single cells identified by DNA (191^Ir and ^193^Ir)、^194^Pt and event length. For downstream analysis, 10,000 cells were randomly selected from each sample. The raw values of FCS files were transformed using the arcsinh function with a cofactor of 5. Phenograph clustering in cytofkit R package was performed on all subsampled sample events and the transformed values of all events were subjected to t-SNE dimension reduction using R-tsne package^32,33^.

### Antibodies

The following antibodies were purchased from Cell Signaling Technology: pRIPK1 (S166) (65746), RIPK1 (3493), pMLKL (S358) (91689), MLKL (14993), pIKKα/β (S176/180) (2697), IKKβ (2370), Sharpin (12541), pp65 (S65) (3033), p65 (8242), pIκBα (S32) (2859), IκBα (4814), pERK1/2 (T202/Υ204) (4370), ERK1/2 (4695), pJNK (T183/Y185) (9251), JNK (9252), HA (3724), cleaved caspase-3 (D175) (9661), caspase-3 (9662). NEMO (sc-8330) was purchased from Santa Cruz Biotechnology. GAPDH (SA30-01) was purchased from HUABIO.

## Data availability

The raw data of sequencing will be available prior to publication. The other source data that support the findings of this study are available from the corresponding author upon reasonable request.

---

**Extended Data Fig. 1.**
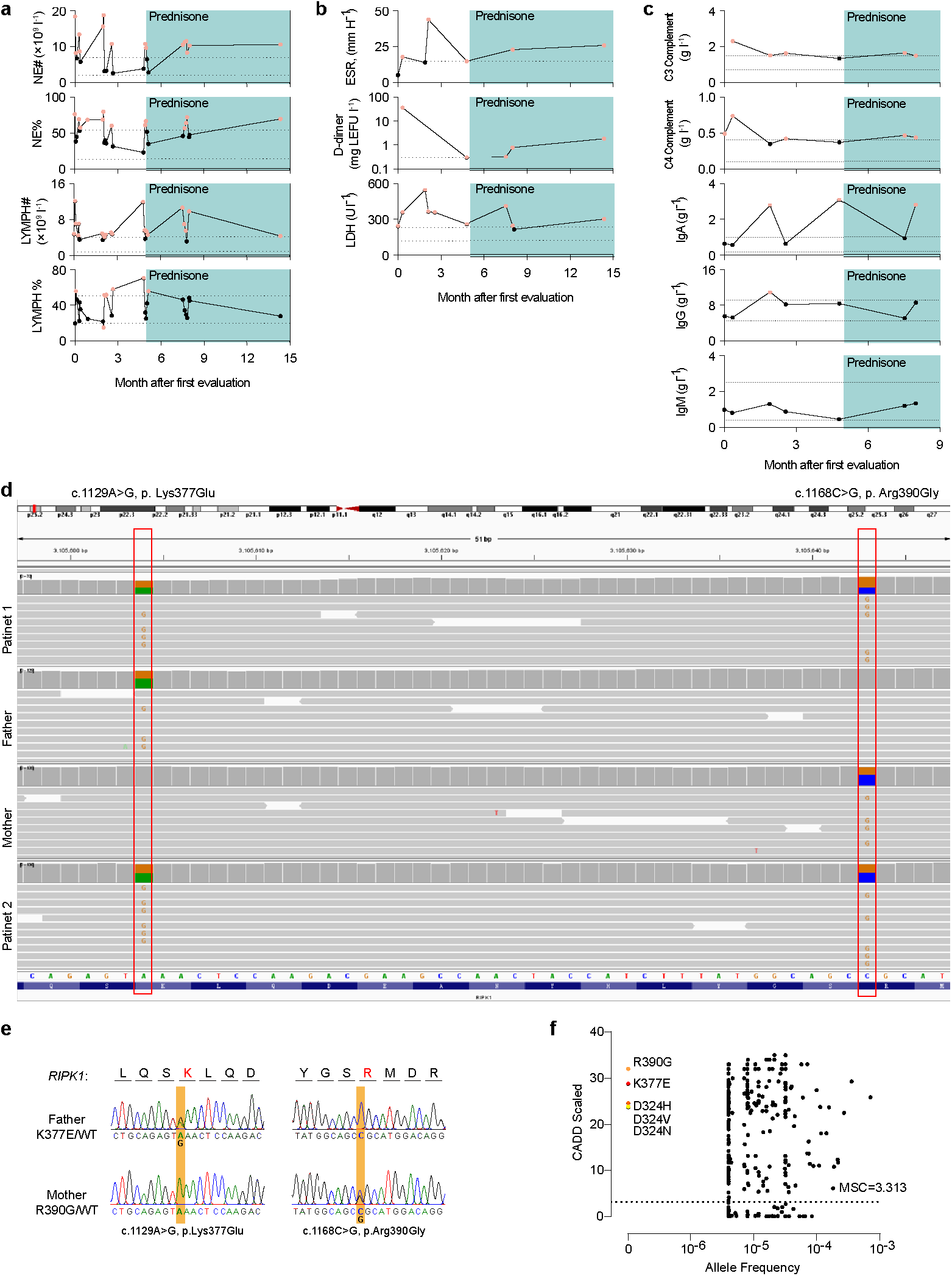
Clinical and genetic information of autoinflammatory patients carrying compound heterozygous GOF mutations in *RIPK1*. **a-c,** Cell counts and proportions of neutrophil (NE#-NE%) and lymphocyte (LYMPH# -LYMPH%) (**a)**, erythrocyte sedimentation rate (ESR), D-dimer, serum lactate dehydrogenases (LDH) (**b)**, serum C3 complement, C4 complement, IgA, IgG, IgM (**c)** measured serially after the first evaluation of P1 and prednisone treatment (cyan shading) started at the age between 0-5. Horizontal dotted lines indicating age-specific high values for ESR, D-dimer or high and low values for the rest tests. Red dots donate the abnormal values. **d,** Whole exome sequencing reads covering *RIPK1* p. Lys377Glu (NM_003804.4: c.1129A>G) and p. Arg390Gly (NM_003804.4: c.1168C>G) variants in two patients and their parents displayed using integrative genomics viewer. **e,** Sanger sequencing chromatograms of *RIPK1* in whole genomic DNA from parents. **f,** Correlating Combined Annotation Dependent Depletion (CADD) scores with minor allele frequencies (MAF) for *RIPK1* K377E, R390G and D324 variants. Mutation significance cutoff (MSC) was show.

**Extended Data Fig. 2.**
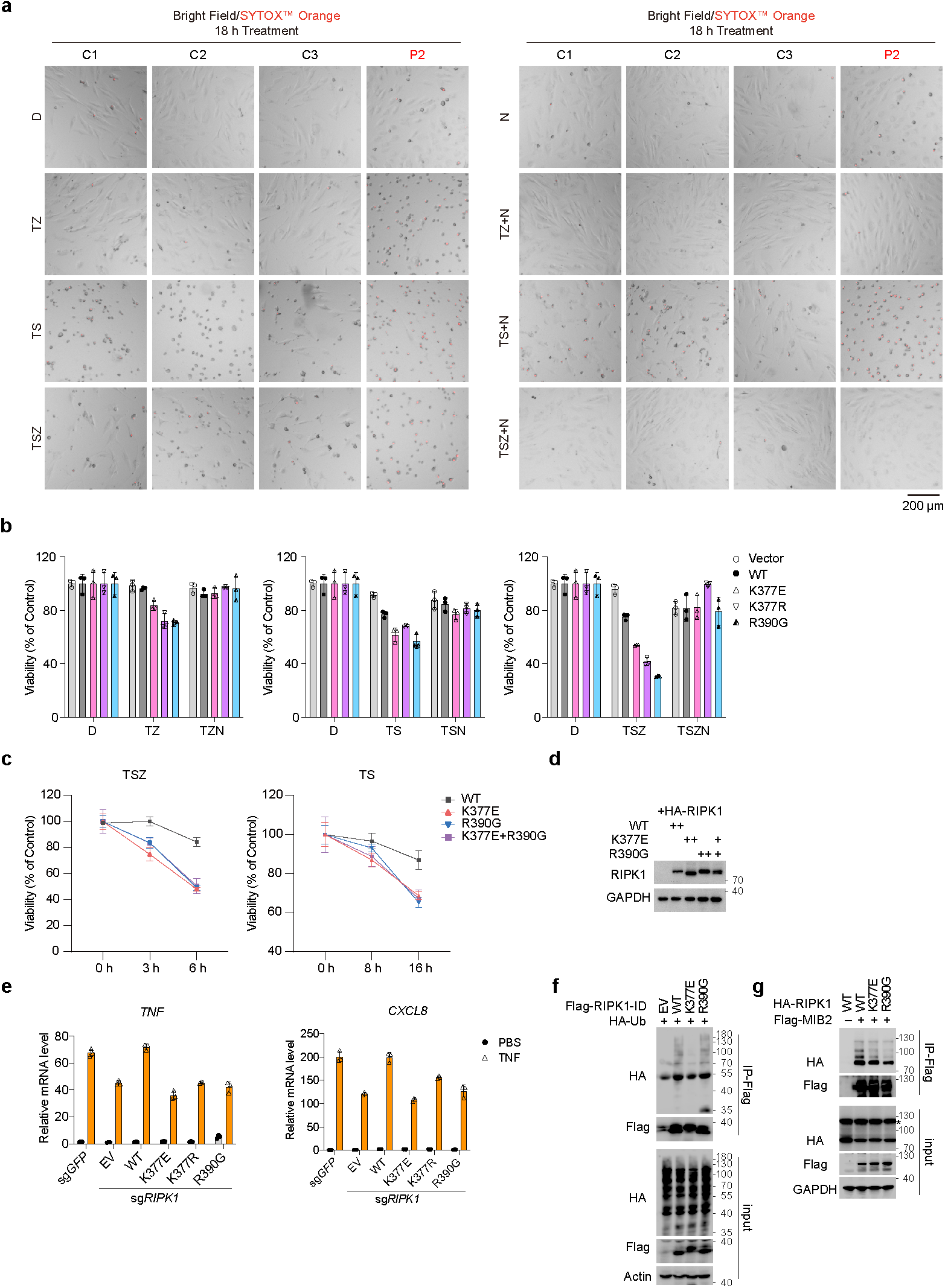
*RIPK1* mutations sensitize cells to TNF-induced cell death. **a,** SV40-HDFs isolated from three unaffected controls (C1-C3) and P2 were treated with T with or without S, Z and N as indicated. Cell death were monitored by imaging of SYTOX™ Orange staining. **b-d,** CellTiter-Glo luminescent cell viability detection of *RIPK1*^-/-^ HT-29 cells complemented with WT or mutated *RIPK1* K377R, K377E, R390G, K377E/R390G under stimulation of TZ, TS, TSZ with or without N as indicated **(b-c)**. Expression levels of WT or mutated RIPK1 were determined with immunoblot **(d)**. **e,** mRNA levels of *TNF*, *CXCL8* in complemented HT-29 cells treated with TNF for 6 hours were determined with qRT-PCR. **f-g,** HEK293T cells were transfected with plasmids encoding full length/intermediate domain of RIPK1 together with MIB2 or Ub as indicated. ID domain of RIPK1 or MIB2 were immunoprecipitated with anti-Flag dynabeads and immunoblotted with indicated antibodies. T denotes 20 ng ml^-1^ TNF; S denotes 250 nM SM-164; Z denotes 25 μM z-VAD-fmk; N denotes 10 μM Nec-1s. Graphs show mean±SD.

**Extended Data Fig. 3.**
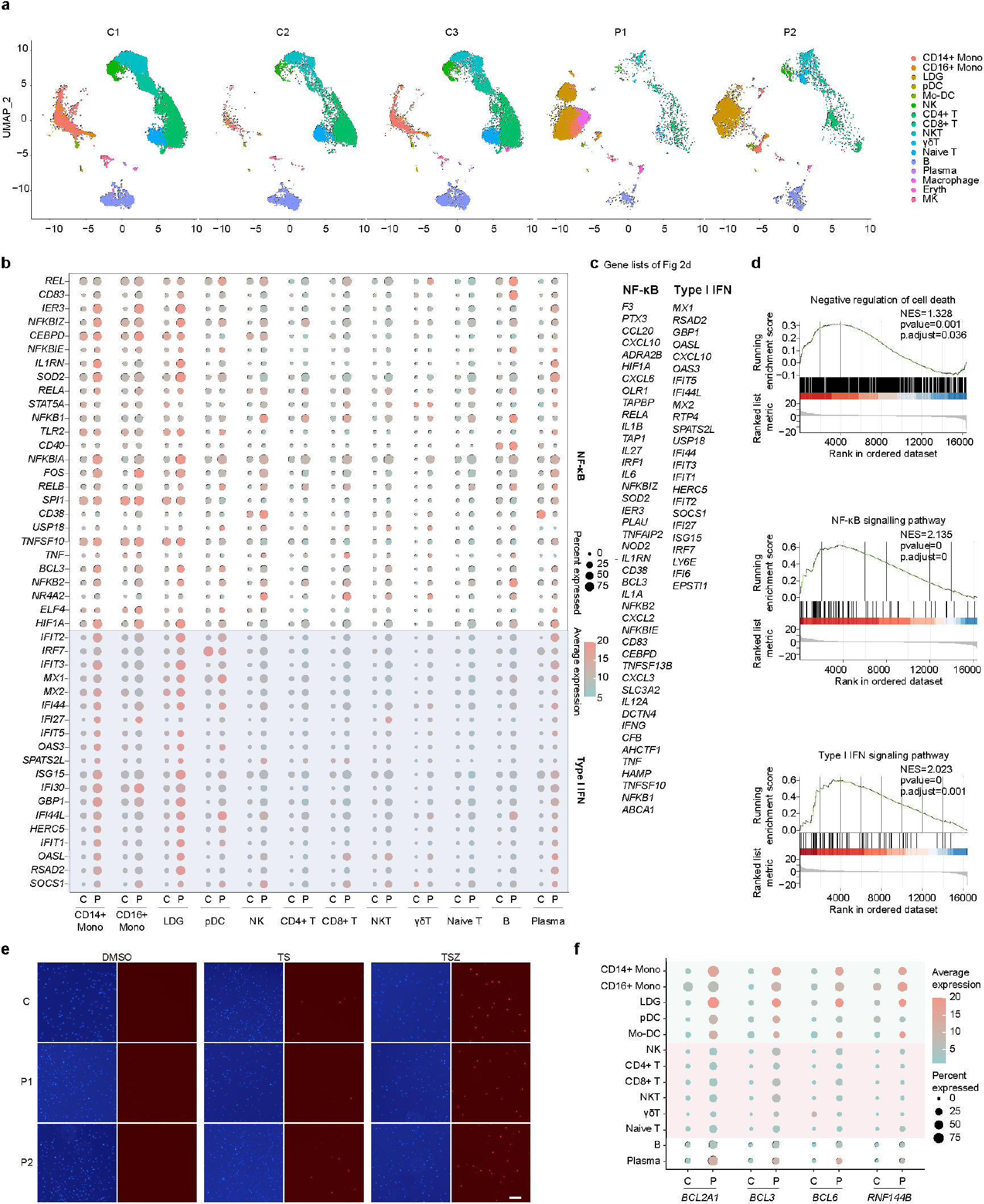
Inflammatory signature in patients’ monocytes. **a,** Uniform manifold approximation and projection (UMAP) visualization of peripheral blood mononuclear cells (PBMCs) from three unaffected controls (C1, n=13925 cells; C2, n=11117 cells; C3, n=12573 cells) and two patients (P1, n=10918 cells; P2, n=8516 cells). **b,** Dot plot showing the expression of representative genes downstream of NF-κB and type Ι IFN pathways in cell subtypes of PBMCs. The size of the circle indicates the percentage of cells positive for target gene expression and the color represents the average expression levels of genes in clusters. **c,** List of gene names in keeping with Fig.2d. **d,** Gene set enrichment analysis (GSEA) plot of negative regulation of cell death (NES=1.328, P<0.01), NF-κB signaling pathway (NES=2.135, P<0.01) and Type I IFN signaling pathway (NES=2.023, P<0.01). NES, normalized enrichment score. **e,** Representative images of dead M1 macrophages in keeping with Fig.2i. **f,** Dot plot of negative regulators of cell death *BCL2A1, BCL3, BCL6, RNF144B* in cell subtypes found in scRNA-seq of PBMCs from three unaffected controls and two patients. The size of the circle indicates the percentage of cells positive for gene expression and the color represents the average expression levels.

**Extended Data Fig. 4.**
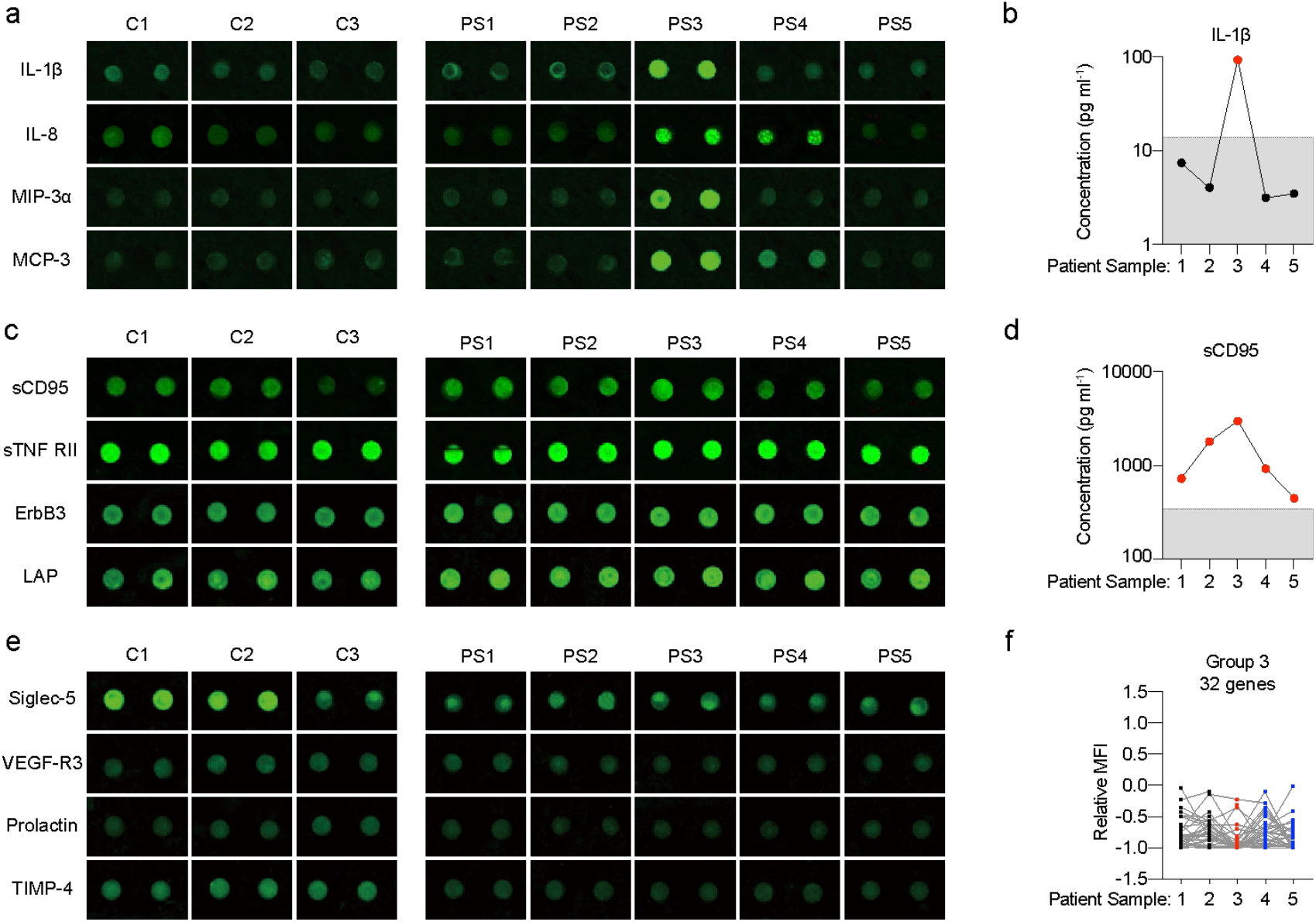
Cytokine array analysis of patient’s serum. Images of representative cytokines in group 1 (**a**), group 2 (**c**) and group 3 (**e-f**) clustered from cytokine array data according to Fig. 2l-n were presented. Typical cytokines were re-validated with ELISA (**b, d**) and integrated relative MFI plots of group 3 cytokines were shown.

**Extended Data Fig. 5.**
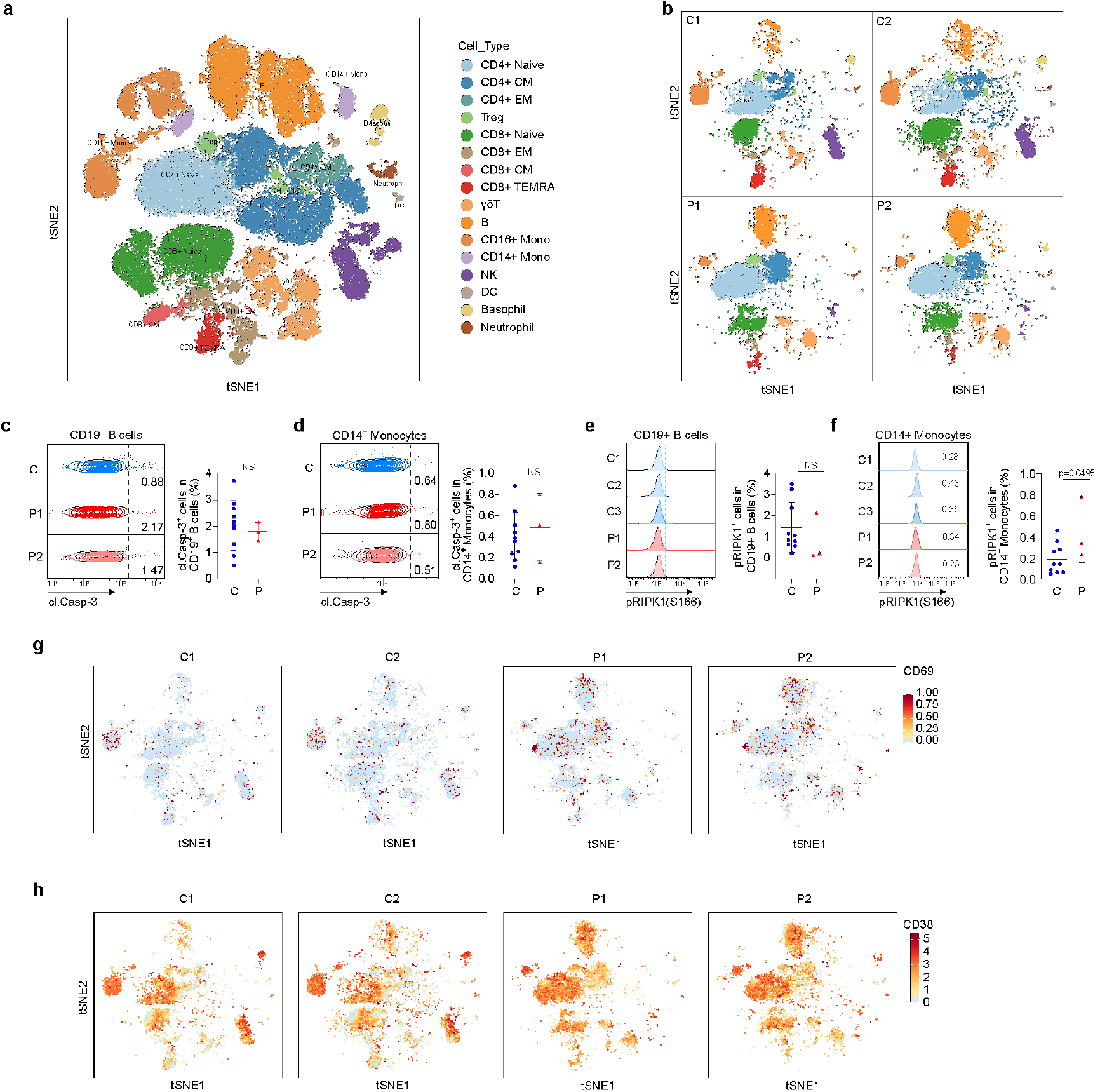
CyTOF profiling of patients’ PBMCs. **a-b,** Integrated t-distributed stochastic neighbor embedding (tSNE) visualization of CyTOF immunophenotyping of PBMCs from four unaffected controls and two patients (**a**). Major 16 cell lineages from all samples were identified through unsupervised clustering with FlowSOM. tSNE plots showing 10000 cells of representative C1, C2, P1 and P2 with color-indicated cell types (**b**). **c-f,** Representative contour plots of cl. Casp-3 (**c-d**) and histograms of pRIPK1(S166) (**e-f**) protein levels in CD19^+^ B cells and CD14^+^ monocytes in freshly isolated PBMCs from unaffected controls (n=10) and patients (n=3 for P1 and P2) (left panel). Summary histograms were shown with mean±SD (right panel). **g-h,** t-distributed stochastic neighbor embedding (tSNE) plot showing the CD69 and CD38 expression in CyTOF analysis. Statistical significance was determined by two-tailed unpaired t-test. ns, *P* > 0.05, \**P*<0.05, \*\**P* < 0.01, \*\*\**P* < 0.001.

**Extended Data Fig.6.**
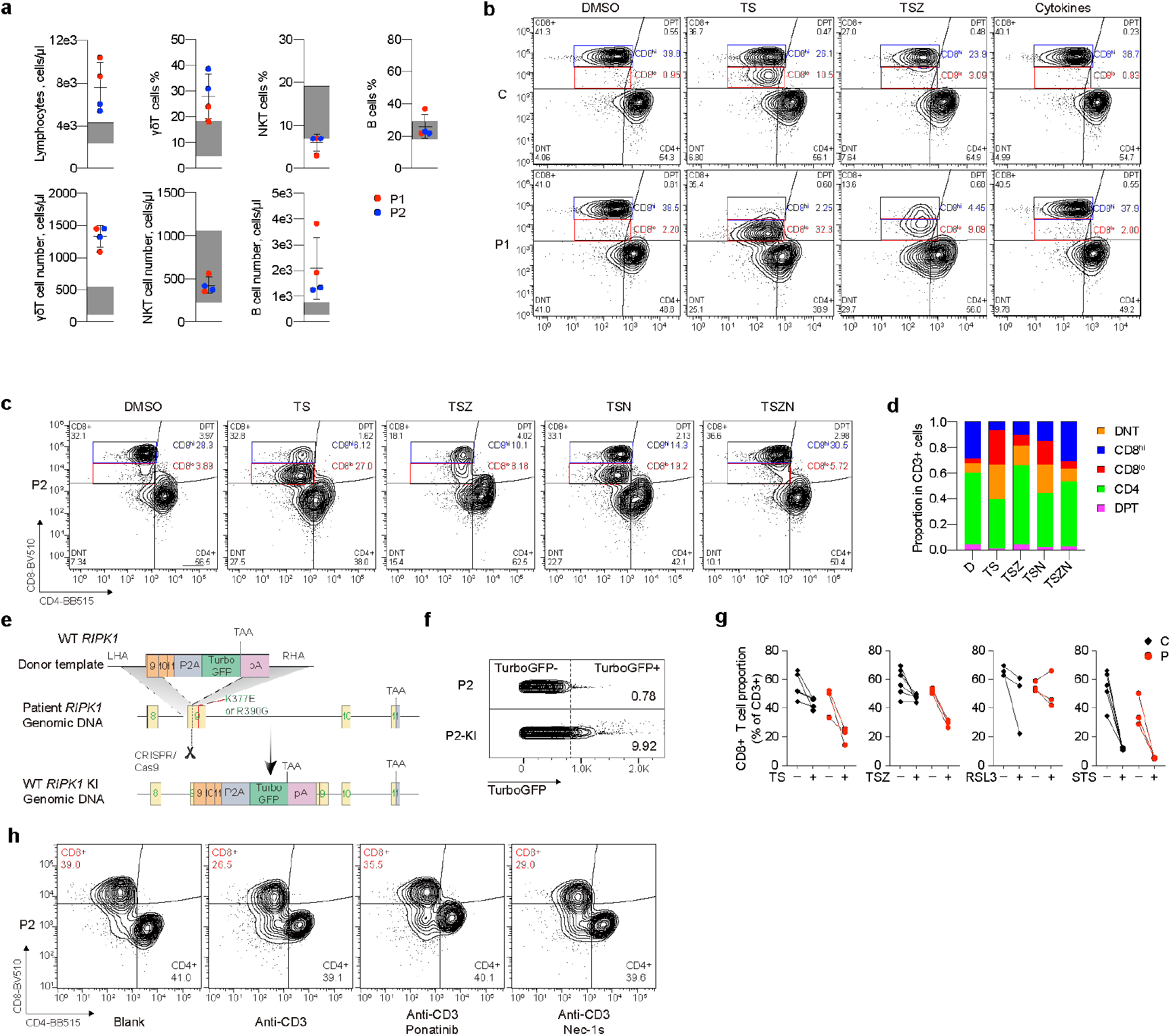
RIPK1-mediated cell death leads to CD8 decrease in T cells. **a,** Percentages and absolute counts of total lymphocytes, γδ T cells, NKT cells and B cells from whole blood of P1 (red dots) and P2 (blue dots) measured by immunophenotyping. **b,** Representative contour plots showing percentages of CD8^hi^ T, CD8^lo^ T, CD4^+^ T, DNT and DPT cells in CD3^+^ T cells isolated from unaffected control and P1 under the stimulation of TS, TSZ and cytokines (combination of IL-1β (10 ng ml^-1^), IL-6 (100 ng ml^-1^), IFNα (100 ng ml^-1^) and IL-8 (100 ng ml^-1^)) as indicated for 6 hours. **c- d,** Representative contour plots (**c**) and summary histogram (**d**) showing percentages of CD8^hi^ T, CD8^lo^ T, CD4^+^ T, DNT and DPT cells in CD3^+^ T cells isolated from P2 under the stimulation of TS, TSZ, TSN and TSZN as indicated for 6 hours. **e,** Schematic of CRISPR/Cas9-mediated *in situ* knock-in of WT *RIPK1* into patients’ *RIPK1* locus. **f,** Representative contour plots of Turbo-GFP^+^ cells percentage indicating WT *RIPK1* knockin efficiency. **g,** Statistical analysis of CD8^+^ T cell proportions in CD3^+^ T cells from unaffected controls (n=3-6) and two patients (n=3 for P1 and P2) under the stimulation of TS, TSZ as well as RSL3 (1 μM) and STS (100 nM) for indicated times. **h,** Representative contour plots showing percentages of CD8^+^ T and CD4^+^ T cells in CD3^+^ T cells isolated from P2 under the stimulation of anti-CD3 antibody (5 μg/ml) with or without Ponatinib (250 nM) and Nec-1s (10 μM) for 24 hours. T denotes 20 ng ml^-1^ TNF; S denotes 250 nM SM-164; Z denotes 25 μM z-VAD-fmk; N denotes 10 μM Nec-1s. Graphs show mean±SD.

**Extended Data Fig. 7.**
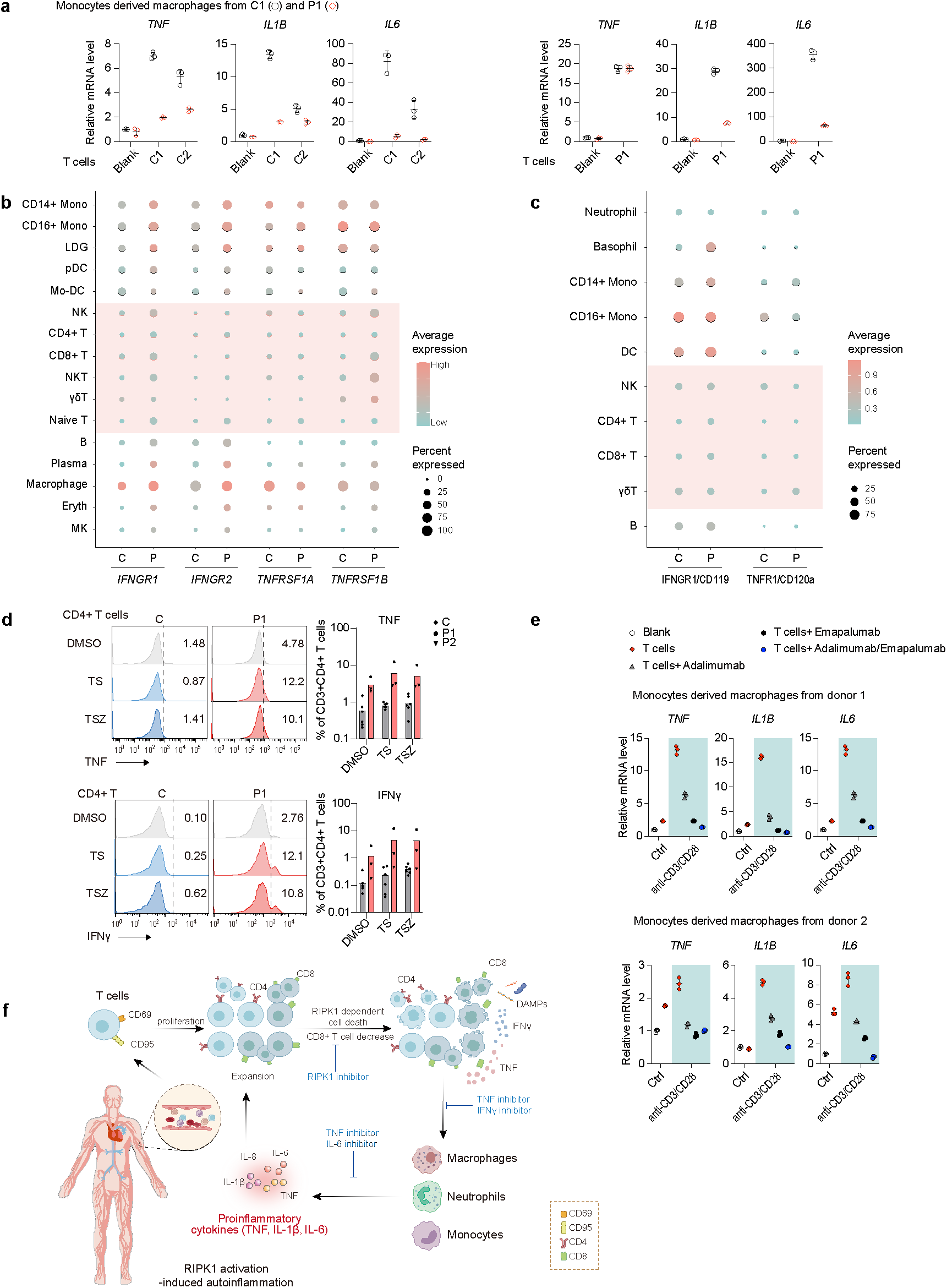
TNF/IFNγ links T-mono axis. **a,** qRT-PCR analysis of *TNF, IL1B* and *IL6* mRNA levels in GM-CSF-activated monocytes derived macrophages (MDMs) co-cultured with or without T cells for 6 hours. GM-CSF activated MDMs generated from C1 (blank dots) or P1 (red dots) were co-cultured with T cells from unaffected controls (C1 and C2, left panel) or P1 (right panel) as indicated. **b-c,** Dot plot showing the mRNA expression of *IFNGR1*, *IFNGR2*, *TNFRSF1A* and *TNFRSF1B* from sc-RNAseq analysis (**b**) or protein levels of IFNGR1/CD119 and TNFR1/CD120a from CyTOF analysis (**c**) in cell subtypes found in PBMCs. The size of the circle indicates the percentages of cells positive for gene expression and the color represents the average expression levels. **d,** Representative histograms and statistical analysis of TNF and IFNγ expression in CD4^+^ T cells in PBMCs isolated from unaffected controls (n=6) and patients (n=3 for P1 and P2) treated with TS and TSZ for 24 hours. **e,** T cells stimulated with or without anti-CD3/CD28 dynabeads for 24 hours prior to co-culture with MDMs generated from healthy donor 1 (upper panel) or donor 2 (bottom panel) in the presence of 10 μg ml^-1^ adalimumab, 10 μg ml^-1^ emapalumab alone or in combination for 6 hours. *TNF, IL1B* and *IL6* mRNA levels of MDMs were analyzed by qRT-PCR. **f,** Working model: RIPK1 activation-dependent CD8^+^ T cell death-triggered inflammatory response of myeloid cells cause SAID. Dots represent each image and graphs show mean±SD.

**Extended Data Fig. 8.**
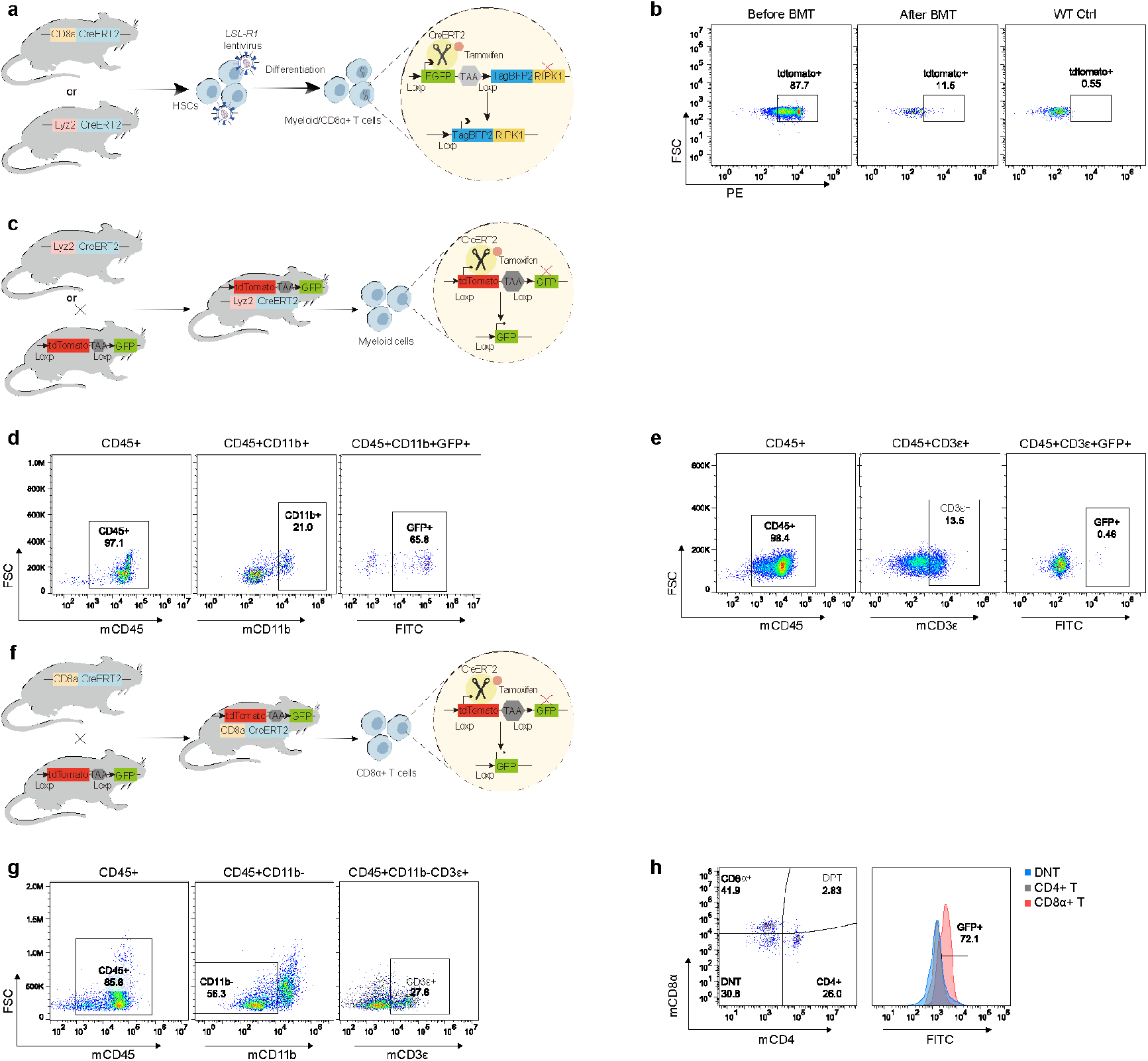
Specificity and efficiency of conditional expression system. **a,** Schematic representation depicting conditional human RIPK1 overexpression system in murine HSC used in this study. **b,** Representative contour plots showing the bone marrow transplantation efficiency. **c-h,** Schematic representation depicting the generation of double transgenic mice used to evaluate the specificity and efficiency of *Lyz2*-CreERT2 (**c**) and *Cd8a*-CreERT2 (**f**). Representative contour plots showing percentage of GFP^+^ cells in myeloid (**d**) and T lymphoid lineage(**e**), as well as in T cell subsets (**g, h**) from the whole blood of *Lyz2*-CreERT2::*Rosa26*-mTmG and *Cd8a*::*Rosa26*-mTmG mice with tamoxifen induction.

**Extended Data Fig. 9.**
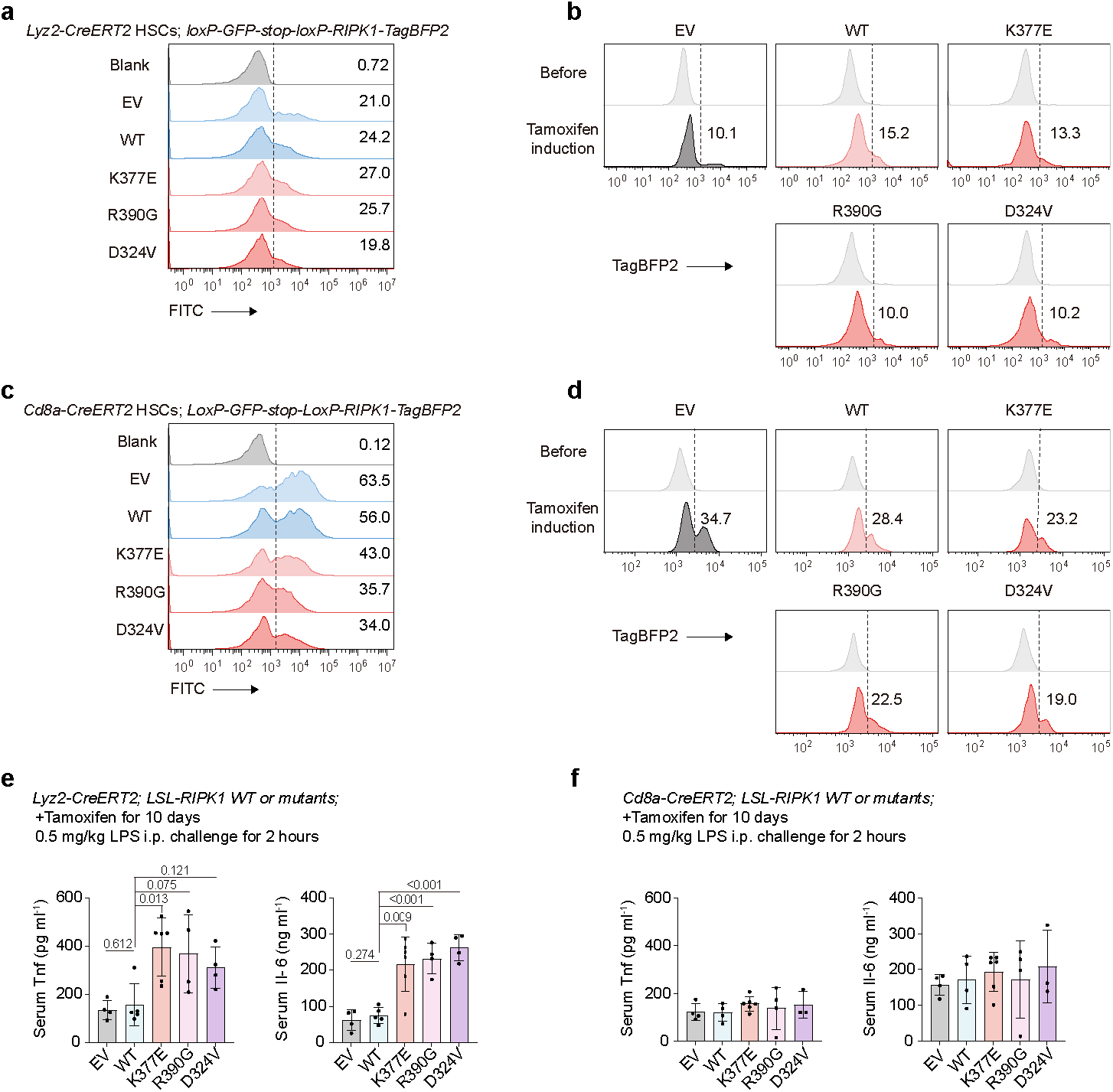
Efficiency of conditionally induced overexpression of RIPK1 in mice. **a-c,** *loxp-GFP-stop-loxp-TagBFP2-P2A-WT/mutated human RIPK1* were lentivirally transduced into HSCs isolated from *Lyz2*-CreERT2 (**a**) or *Cd8a*-CreERT2 donor mice (**c**). Histogram graphs showing expression levels of GFP in HSCs. Whole blood cells isolated from recipient mice with *Lyz2*-CreERT2 (**b**) and *Cd8a*-CreERT2 (**d**) HSCs transplantation before and after tamoxifen induction. Histogram graphs showing expression levels of TagBFP2 in myeloid cells (**b**) and CD8α^+^ T cells (**d**). **e-f,** ELISA measurement of serum Tnf and Il-6 levels in mice specifically overexpressing WT or mutated hRIPK1 in myeloid cells (**e**) or CD8α^+^ T cells (**f**) with 0.5 mg/kg LPS challenge for 2 hours. Statistical significance was determined by two-tailed unpaired t-test.

**Extended Data Fig. 10.**
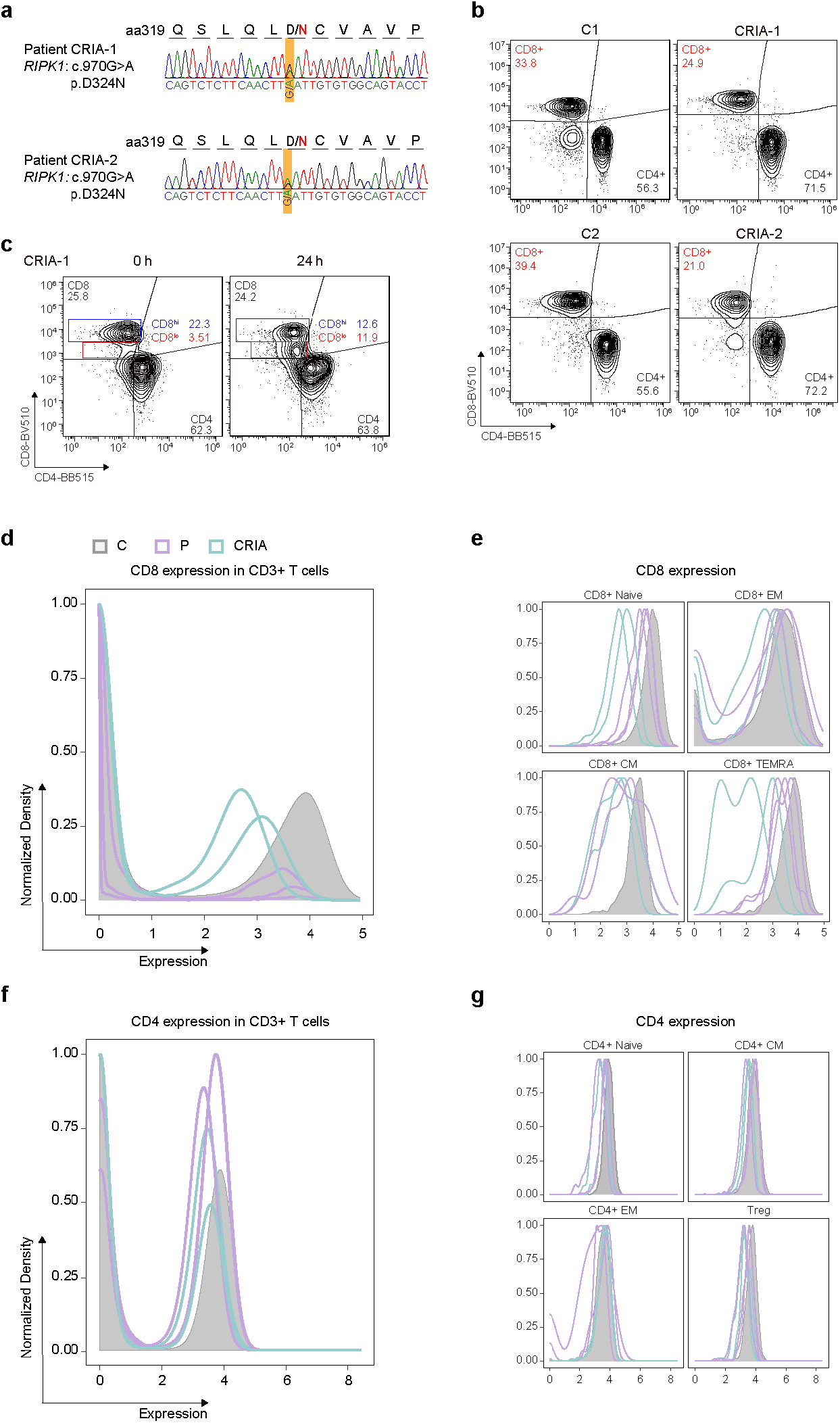
CD8 expression in CRIA patients. **a,** Sanger sequencing chromatograms of *RIPK1* in whole blood genomic DNA from CRIA patients (CRIA-1 and CRIA-2). **b**, Representative contour plots showing percentages of CD4^+^ T and CD8^+^ T cells in CD3^+^ T cells isolated from CRIA patients. **c,** Representative contour plots depicting the percentages of CD4^+^ T, CD8^+^ T, double negative T (DNT) and double positive T (DPT) in CD3^+^ T cells gated from PBMCs of CRIA-1 culturing *in vitro* for 24 hours. **d-g,** Contour plots showing the expression of CD8 (**d**) or CD4 (**f**) in CD3^+^ T cells, CD8^+^ (**e**) and CD4^+^ (**g**) T cell subsets of four unaffected controls, two patients (P1 and P2) and two CRIA patients. The protein expression was determined using CyTOF.

